# Personalized Thalamic Electrical Stimulation for Focal Epilepsy

**DOI:** 10.1101/2024.10.04.24314797

**Authors:** Arianna Damiani, Sirisha Nouduri, Jonathan C. Ho, Steven Salazar, Aude Jegou, Eliza Reedy, Naoki Ikegaya, Sridevi Sarma, Thandar Aung, Elvira Pirondini, Jorge A. Gonzalez-Martinez

## Abstract

Targeted electrical stimulation to specific thalamic regions offers a therapeutic approach for patients with refractory focal and generalized epilepsy who are not candidates for resective surgery. However, clinical outcome varies significantly, in particular for focal epilepsy, influenced by several factors, notably the precise anatomical and functional alignment between cortical regions generating epileptic discharges and the targeted thalamic stimulation sites.

Here we hypothesized that targeting thalamic nuclei with precise anatomical and functional connections to epileptic cortical areas (an approach that we refer to as hodological matching) could enhance neuromodulatory effects on focal epileptic discharges. To investigate this, we examined three thalamic subnuclei (pulvinar nucleus, anterior nucleus, and ventral intermediate nucleus/ventral oral posterior nuclei) in 32 focal epilepsy patients.

Specifically, we first identified hodologically organized thalamocortical fibers connecting these nuclei to individual seizure onset zones (SOZs), combining neuroimaging and electrophysiological techniques. Further, analysis of 216 spontaneous seizures revealed the critical role of matched thalamic nuclei in seizure development and termination. Importantly, electrical stimulation of hodologically-matched thalamic nuclei immediately suppressed intracortical interictal epileptiform discharges, contrasting with ineffective outcomes from stimulation of unmatched targets. Finally, we retrospectively evaluated 7 patients with a chronic hodologically-matched neurostimulation system, which led to a clinically relevant reduction in seizure frequency (median reduction 86.5%), that outstands the current clinical practice of unmatched targets (39%).

Our results underscore the potential of hodological thalamic targeting to modulate epileptiform activity in specific cortical regions, highlighting the promise of precision medicine in thalamic neuromodulation for focal refractory epilepsy.

## Introduction

Over 50 million people worldwide suffer from epilepsy, a neurological disorder characterized by recurrent seizures, stemming from synchronized and excessive paroxysmal electrical activity of neurons^1,2^. In most of the cases (>60%), said seizures originate and develop in restricted and focal cortical regions of the brain^3^, condition clinically referred to as focal epilepsy. Conversely, in generalized epilepsy, pathological brain activity stems from widespread areas of the brain, affecting both hemispheres^4,5^. Independently of the focal or generalized onset, about 30% of epilepsy patients live with intractable disabling seizures^6^, which cannot be controlled with medical therapy. The current standard of care for medically refractory epilepsy (MRE) relies on the resection of the seizure focus, i.e., the brain region associated with earliest electrophysiological changes during a seizure event^7^. However, if said cortical areas are not amenable for safe removal (or disconnection) because of overlap with eloquent cortices, or extensive seizure foci, or a diagnosis of generalized epilepsy, neuromodulation approaches such as thalamic electrical stimulation might be an alternative^7–11^. Recent studies have propelled thalamic stimulation for epilepsy to the forefront of neuroscience, underscoring its emerging role as a pivotal therapeutic strategy in the field^12^.

The thalamus is a deep central structure crucial for its multifaceted functions and intricate organization. Indeed, through extensive connections with the cortex and other subcortical and peripheral structures, it is integral to a wide array of functions including motor and sensory processing, consciousness, and sleep^13–16^. This diverse functionality is attributed to the thalamus’s varied nuclear architecture and its intimate relationship with cortical areas^17^. Importantly, since the early 1950s the thalamus was demonstrated to synchronize widespread cortical activity, hence contributing to the generation of pathological epileptic discharges^18^. Indeed, altered thalamocortical synchrony was extensively observed during focal seizures and associated loss of consciousness^19^. Furthermore, structural connectivity variations was reported between the thalamus and the cortex in patients with generalized epilepsy^20^.

Given these cortico-subcortical dynamics and epileptogenic interactions, thalamic electrical stimulation has been proposed since the early 80s, with indications both in focal^9,21^ and generalized epilepsy^8^. In this regard, thalamic sites characterized by diffuse connectivity as the centromedian nucleus (CM), have been hypothesized to benefit generalized seizures, which originate and develop through widespread neuronal networks. Indeed, in a recent clinical trial, Cukiert and colleagues reported a significant seizure reduction (>50%) with CM electrical stimulation in 90% of tested patients diagnosed with generalized epilepsy^22^. CM stimulation, instead, has been less effective in patients with focal MRE^23^. For this reason, thalamic nuclei with less diffuse connectivity, and mostly the anterior nucleus (ANT), have been explored as a possible target for neuromodulation in focal epilepsy. Already in the early 80s, Cooper et al. showed a significant clinical control of seizures in 4 out of 6 patients with stimulation of the ANT^24^. Since this seminal work, multiple studies have investigated the efficacy of ANT stimulation to treat focal epilepsy^9,21,25^, including the pivotal SANTE trial that achieved 75% median seizure reduction at 7 years follow up^26^. Thanks to these works, ANT stimulation for focal epilepsy received FDA approval in 2018. However, clinical outcomes are highly variable depending on the anatomical location of the seizure onset. Specifically, the most well-documented efficacy was observed in focal onset seizures occurring in the temporal and frontal lobe (78% and 86% seizure frequency reduction), whereas effects were limited for other cortical seizure onset zones (SOZs) (39%)^26^. This might be due to the ANT’s unique anatomical connectivity to temporal and frontal areas^27,28^.

Exploration in other areas of the thalamus further suggests a possible link between thalamocortical anatomical connectivity and neuromodulation efficacy. Indeed, the pulvinar nucleus (PUL) of the thalamus, which has high connectivity towards the temporal and occipito-parietal lobes and present different neural firing patterns during ictal progression and termination in patients with temporal lobe epilepsy^29–32^, has been shown to be potentially effective for posterior quadrant epilepsy^33,34^. Similarly, the ventrolateral portion of the thalamus (ventral intermediate /ventral oral posterior nuclei, VIM/VOP)^35^ and the cerebellum^36^, which present high structural connectivity to motor cortical areas, were tested, with positive outcome, for epilepsies organized in the agranular pre-central neocortical areas (rolandic epilepsy), which is particularly challenging for resective surgeries due to the high risks of motor deficits.

However, these studies remain isolated investigations leading to a lack of consensus on which target is optimal for focal epilepsy and clinical indication on how to approach the heterogeneity of SOZs, with limited clinical outcome. Based on prior evidence, here we argue that, for focal epilepsy, thalamic electrical stimulation should match the hodology of the thalamocortical pathway to enhance therapeutic outcome. In other words, targeting electrical stimulation to the thalamic subnucleus with preferential anatomical and functional connectivity to the SOZ (an approach that we refer to as *hodological matching*) will improve the efficacy of thalamic stimulation for the treatment of refractory focal epilepsy.

To test this hypothesis, we studied the anatomical and functional properties of neural activity of 32 patients with focal MRE. We focused on the study of three thalamic nuclei that exhibit more specific and less diffuse thalamocortical projections as compared to the CM: the ANT, the PUL, and the VIM/VOP. First, we defined a hodological map of the thalamocortical fibers from said thalamic nuclei towards the cortex using imaging and electrophysiology. In this way, we identified the cortical areas that match each nucleus, i.e., receiving major projections. We then characterized the functional involvement of these thalamic nuclei to ictal events occurring both in hodologically-matching and unmatching SOZs. Furthermore, we showed that electrical stimulation of only hodologically-matched thalamic nuclei immediately suppressed pathological discharges i.e., interictal spikes, in the epileptogenic cortex. Finally, we retrospectively verified that such effects led to significant clinical improvement by reducing seizure frequency in 7 patients with a chronic hodologically-matched thalamic implant. Importantly, our hodology-based approach achieved a median reduction in seizure frequency of 86.5%, which dramatically outperforms unmatched thalamic targeting reported by literature (39%).

In summary, our approach, i.e., targeting electrical stimulation to match the hodology of thalamocortical connections, holds significant potential to improve the therapeutic efficacy of thalamic electrical stimulation in the management focal refractory epilepsy. This strategy introduces innovative avenues for clinical targeting and therapeutic intervention, while challenging the prevailing paradigm that adopts a “one-nucleus-fits-all” SOZs.

## Methods

### Trial and participants information

All experimental protocols were approved by the University of Pittsburgh Institutional Review Boards (IRB) (protocol STUDY20070113 and STUDY21020058). All participants included in this study were diagnosed with drug-refractory epilepsy and underwent stereoelectroencephalography (SEEG) implantation at the University of Pittsburgh Medical Center as a part of their standard clinical care. They were hospitalized in the epilepsy monitoring unit (EMU) from January 2021 to May 2024, for 5.04±6.28 days (mean±SD). All patients received a comprehensive neurological assessment, neuropsychological testing, routine MRI and CT and SEEG implantation. Informed consent was obtained from all patients or legal representatives. In order to study the anatomical and functional properties of the thalamus and the epileptic cortex via SEEG monitoring, we retrospectively included patients (S01-S26) in the present study if: 1) at least one SEEG electrode contact was in the PUL, ANT or VIM/VOP, and 2) at least one of the electrophysiological recordings analyzed in the study was performed (minimum 2 spontaneous seizures, thalamocortical evoked potentials, HDFT, thalamic stimulation). Exclusion criteria encompassed refusal in participating in the study and SEEG implantation complication (severe intracranial hemorrhage). We additionally included 7 participants (S16, S27-S32) that received a chronic thalamic stimulation system as part of their clinical care to evaluate long-term efficacy of matched thalamic stimulation.

A comprehensive overview of the patients’ demographics and epilepsy categorization, summary of the experiments, and a summary of the data collected for each patient are reported in **Supplementary Table 1&2**. Overall, we included 26 SEEG participants (20 males, 6 females) and 7 chronically implanted patients (S16, S27-S32), of age 34.8±11.3 (mean±SD). The age at seizure onset was 14.3±13.6 years-old, while the duration of the epilepsy disease was 20.4±13.1 years (mean±SD). Additional information can be found in the **Supplementary Tables 1, 2, 4**.

### SEEG implantation and localization

The number and location of SEEG electrodes implanted was pre-operatively planned individually for each patient as part of their clinical care and was not influenced by this study. The implant procedure was performed with robotic stereotactic guidance (ROSA, Zimmer-Biomet, Warsaw, IN, USA), applying bone fiducial registration with accuracy <0.5 mm). Overall, we implanted a total of 5214 contacts (Microdeep® SEEG Electrodes, 12 to 18 channels, DIXI Medical, Marchaux-Chaudefontaine, France), with an average per patient of 200.53±27.87. To target the thalamic nuclei of interest, we used the following stereotactic coordinates: 1) for PUL, in relation to the AC/PC reformatted planes, and having the PC (posterior commissure) as the reference point, coordinates are X: 2-10 mm lateral to the midline plane; Y: 0 to −6 mm posterior to PC; Z: 0 to −6 mm below the AC/PC defined horizontal plane; 2) for ANT, in relation to the AC/PC reformatted planes, and having the MC (mid commissural point) as the reference point, coordinates are X: 6-8 mm lateral to the midline plane; Y: 6 to 7 mm anterior to the MC; Z: 2 to 4 mm above the AC/PC defined horizontal plane; 3) for VIM/VOP, lateral (from about 5 to about 15 mm lateral to the AC/PC line); anterior/posterior (from about 2 to about 10 mm anterior to PC); and dorsal/ventral (from about +1 to about −2 mm from the AC/PC plane.

To reconstruct the position of the SEEG electrode, we used CURRY software (Compumedics NeuroScan, Hamburg, Germany). First, we marked key reference points on each pre-implantation MRI T1 scan (namely the AC, PC and the nasion). These images were then co-registered with post-implantation CT, and a clinical team member manually identified and labeled all the SEEG contacts from the CT. All the contact locations of the subject MRI were visually inspected and confirmed by a certified neurosurgeon (JGM).

For all subject-analysis (**Figure 1b**), the electrode coordinates in the subject space MRI were then translated to coordinates in MNI space for each patient. To confirm the contact location within the thalamic nuclei of interest we used The Human Motor Thalamus Atlas^37^. The assessment of SOZ contacts (as well as non-SOZ contacts used in the following analysis) was confirmed by an experienced neurosurgeon (JGM) (**Supplementary Table 3**).

**Figure 1:**
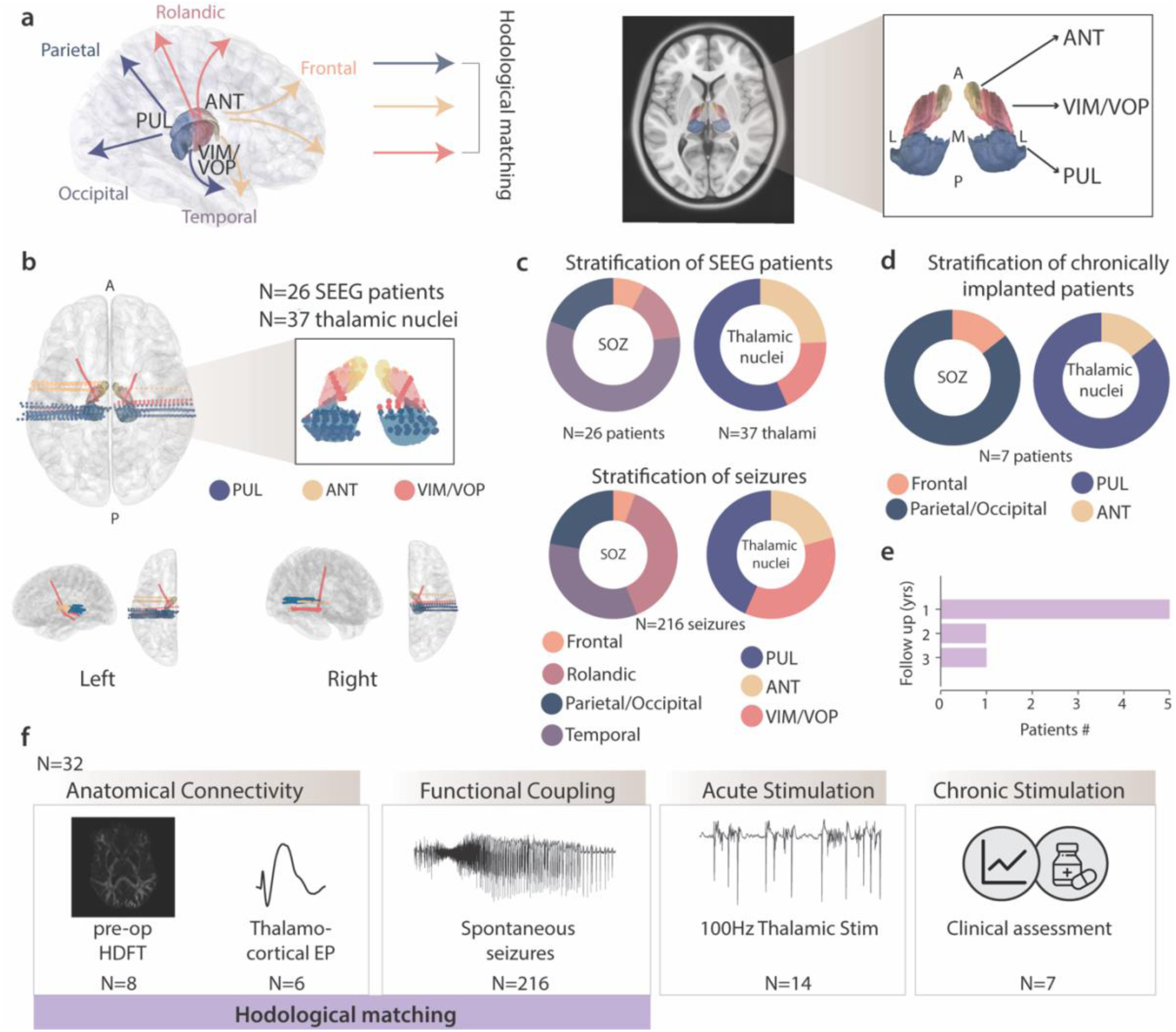
Study design and patient demographics. (**a)** *Left:* Schematic of hodological matching of PUL, ANT and VIM/VOP to specific cortical anatomical regions. *Right:* anatomical localization of PUL, ANT and VIM/VOP nuclei on MNI space average T1-weighted MRI with axial view and magnification of the thalamic nuclei. Thalamic parcellation is atlas-guided^37^ (A=anterior, P=posterior, M=medial, L=lateral). (**b**) *Top right:* axial view of reconstruction of SEEG electrodes targeting thalamic nuclei of interest. *Top left:* magnification of the contacts located in the PUL (blue), ANT (yellow) and VIM/VOP (red). *Bottom:* side and top view of the left and right hemisphere SEEG thalamic implantations. (**c**) Stratification of SEEG patients (S01-S26). *Top Left:* pie chart of the lobar location of the SOZ. *Top Right:* pie chart of 37 thalamic SEEG contacts in PUL, ANT and VIM/VOP. A higher number of thalamic contacts than patients implies that some patients had more than one nucleus of interest implanted. *Bottom:* Stratification of the 216 recorded spontaneous seizures according to the lobar location of different SOZs (left) and the respective thalamic coverage (right). A different distribution from the top panel implies a different number of seizures for each patient, as reported in Supplementary Table 2. (**d**) *Top:* Stratification of 7 chronically implanted patients (S16, S27-32) according to the lobar location of the SOZ (left) and implanted thalamic nucleus (right). (**e**) histogram of follow-up years of chronically implanted patients. (**f**) Design and experiments of the study. Patients received SEEG implantation and were stratified according to their SOZ and thalamic coverage. *Bottom, left to right*: we investigated the anatomical (imaging and electrophysiology), and functional coupling during seizures involving the thalamus, hence defining hodologically matching and unmatching nuclei. We then tested the effect of acute thalamic stimulation and finally the clinical outcome following chronic thalamic stimulation.

### SEEG recording and stimulation

SEEG data were recorded with Natus Quantum System EEG diagnostic and monitoring system (Natus, Pleasanton, A, USA), with a sampling rate of 2048 Hz (for S26 the sampling rate was 1024 Hz) during the extra-operative monitoring at the Epilepsy Monitoring Unit of the University of Pittsburgh Medical Center (Presbyterian Hospital). For all analysis, we applied SEEG bipolar montage to increase spatial selectivity and reduce noise levels. To deliver electrical stimulation in the thalamic contacts, we used the Nicolet Cortical Stimulator (Natus, Pleasanton, A, USA), which delivers biphasic pulses up to 100 Hz (amplitude was set to 1-3 mA and pulse duration to 60-300 μs for all patients) with bipolar electrode configuration.

### High-Definition Fiber Tracking

To estimate anatomical projections from the thalamus to various cortical areas, we first performed High-Definition Fiber Tracking (HDFT) of diffusion MRI data. The diffusion images were acquired on a SIEMENS Prisma Fit scanner using a diffusion sequence (2mm isotropic resolution, TE/TR= 99.2 ms/2490 ms, 257 diffusion sampling with maximum b-value 4010 s/mm²). Diffusion tensor estimation and tractography were performed using DSI studio (http://dsi-studio.labsolver.org). The accuracy of b-table orientation was examined by comparing fiber orientations with those of a population-averaged template. The tensor metrics were calculated using DWI with b-value lower than 1750 s/mm². For fiber tracking, we used a tracking threshold of 0, angular threshold of 0, and a step size of 0 mm. We utilized seed regions in thalamus to create white matter tracts to regions of interest in the cortex. Seed regions were selected in ANT, VIM/VOP, PUL, and CM based on the extended Human Connectome Project multimodal parcellation atlas. Regions of interest were selected in the frontal lobe, rolandic area, parietal lobe, occipital lobe, and temporal lobe. Tracks with lengths shorter than 30 mm or longer than 1000 mm were discarded. A total of 10,000 tracts were placed. Topology informed pruning was applied to the tractography with 2 interactions to remove false connections. We then quantified the volume of white matter tracts projecting from each thalamic nucleus to each cortical area. We normalized the volume of each white matter projection by the total volume projections from each thalamic nucleus (**Figure 2a-b, Extended Data Figure 1 a-b**).

**Figure 2:**
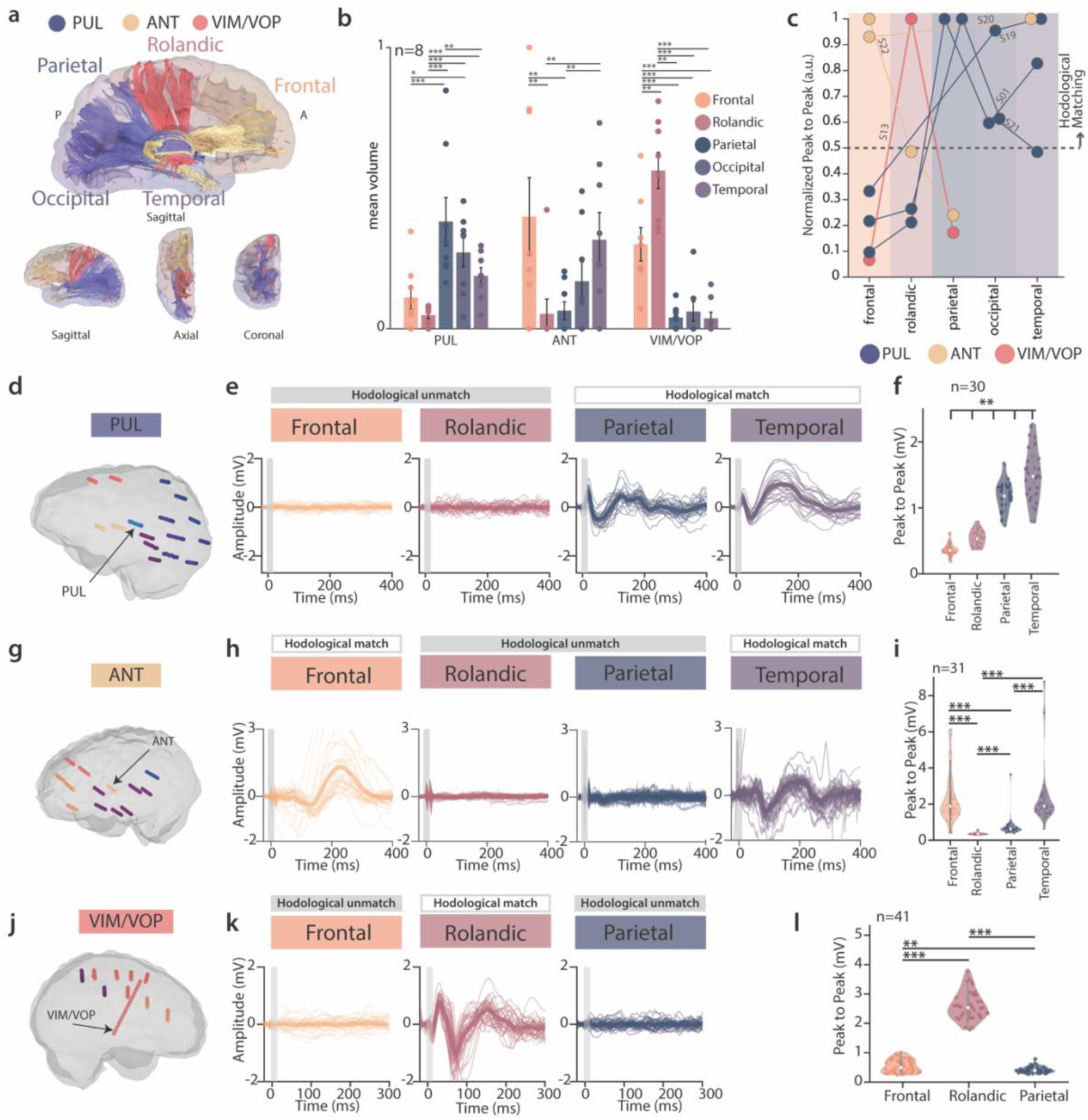
Anatomical connectivity of PUL, ANT and VIM/VOP via neuroimaging and electrophysiology. (**a**) Representative example of high-definition fiber tracking (HDFT) from PUL, ANT and VIM/VOP nuclei (S01). (**b**) Volume of thalamocortical projections (mean ± standard error, n= 8 patients) from each nucleus to each cortical lobe normalized by the total volume of fibers projecting from each nucleus (mean ± standard error). (**c**) Group-level analysis of thalamocortical EPs. For each patient, the mean peak to peak amplitude is calculated across the contacts of a representative electrode for each cortical lobe. The normalized peak to peak amplitude of EP is color coded according to the stimulating nucleus evoking the response (blue for PUL, yellow for ANT, red for VIM/VOP). (**d, g, j**) Anatomical localization of SEEG implantation of representative patients (S01, S22, S13) that were tested for EPs with stimulation of PUL, ANT and VIM/VOP respectively. Each electrode is color-coded according to the brain lobe, and thalamic electrodes are indicated by an arrow. (**e, h, k**) Raw traces of stimulation triggered averages in different brain lobes from PUL, ANT and VIM/VOP respectively. Specific anatomical regions are: frontal operculum (pars opercularis), supplementary motor area, angular gyrus, inferior temporal sulcus (panel e, left to right); superior frontal sulcus, motor cingulate, posterior cingulate, posterior uncus (panel H, left to right); anterior cingulate, supplementary motor area, precuneus (panel K, left to right); (**f, i, l**) Violin plot of peak to peak amplitude of EPs in different brain lobes for PUL, ANT and VIM/VOP respectively. For all panels, statistical significance was assessed with two-tail bootstrapping with Bonferroni correction: p<0.05 (*), p<0.01 (**), p<0.001(***).

### Thalamocortical evoked potentials

In order to confirm the anatomical organization of thalamocortical fibers with electrophysiological techniques, thalamocortical evoked potentials were elicited by thalamic stimulation of PUL, ANT and VIM/VOP (frequency=1 Hz, amplitude=1-3 mA, pulse width=300 μs) and recorded from all available SEEG channels with Natus recording system. SEEG intracerebral data from all cortical channels were filtered with a band-pass Butterworth 2nd order filter (1-1000 Hz) and DC offset was removed from all recordings. From each stimulation pulse, we extracted 520 ms epochs (20 ms prior and 500 ms after the stimulus) and computed stimulation triggered averages. Each epoch was baseline corrected with the pre-stimulus interval. We calculated the peak-to-peak amplitude of evoked potentials in each epoch as the difference between the maximum and minimum voltage value observed within the first 400 ms from the stimulus. We performed this analysis for all available channels that presented sufficient signal to noise ratio, that were located in the regions of interest for this study (not in thalamus) and that were not located in the white matter (**Figure 2d-l, Extended Data Figure 2**).

### Acquisition and analysis of spontaneous seizures

#### Spontaneous seizures acquisition

During the extra-operative monitoring, we recorded 216 spontaneous seizures from 23 patients. The remaining 3 (S14, S16, S19) patients either presented technical problems that invalidated the inclusion of the seizures or did not present any spontaneous events during the hospitalization. For our analysis, we collected recordings from 2 minutes before the seizure onset up to 30 seconds after the seizure termination. These time events (onset and termination) were manually marked for each event by certified medical personnel. We used such markers to define 5 epochs of interested, used in subsequent analysis: 1) baseline (corresponding to 20 seconds occurring two minutes before the seizure onset); 2) seizure initiation (including the 10 seconds preceding the first appearance of a tonic discharge in the SOZ contacts); 3) seizure termination (including the last 10 seconds of discharge); 4) middle of seizure (as the epoch between the previous two intervals 5) total seizure duration (defined as the interval between onset and termination).

#### Non-linear correlation analysis

For each recorded seizure, we computed the h2 non-linear correlation coefficient, as implemented in Anywave^38^ for each pair of thalamus-SOZ bipolar contacts. The h2 coefficient between two signals is a time-resolved measure of their non-linear dependence. The coefficient was computed over a 2 s time window sliding by steps of 1 s. We computed the h2 across the full SEEG spectrum (1-500 Hz), as well as separate frequency bands: delta (1-4 Hz), theta (4-8 Hz), alpha (8-15 Hz), beta (15-30 Hz), gamma (30-45 Hz), gamma2 (55-90 Hz), ripple (80-250 Hz), fast ripple (250-500 Hz). For each time interval of interest (seizure duration, seizure initiation, middle of seizure, seizure termination), we computed the mean h2 (**Figure 3 b, e, f**).

**Figure 3:**
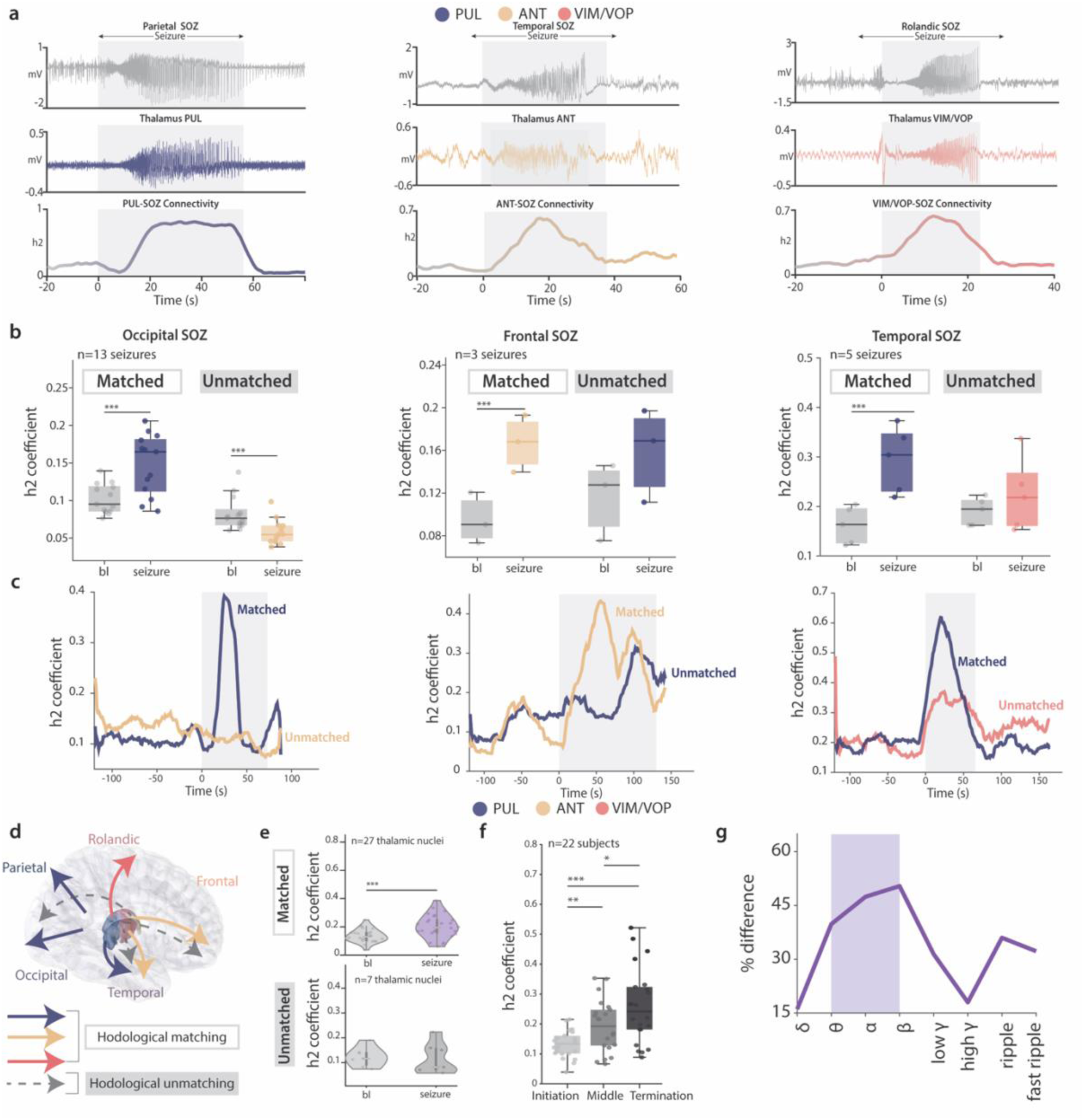
Functional coupling of the PUL, ANT and VIM/VOP nuclei with SOZ during seizures. (**a**) Representative examples of thalamocortical connectivity profiles of PUL (right, S03), ANT (middle, S24) and VIM/VOP (left, S26) with matched SOZ. For all nuclei, the top panel shows the cortical recording in the contacts of the SOZ, the middle panel shows the thalamic recording, and the bottom panel shows the h2 correlation coefficient. The gray shaded area includes the seizure duration (from onset to termination). (**b**) Boxplot of the mean h2 correlation for each seizure in matched and unmatched thalami, compared to mean h2 correlation during a baseline epoch preceding the seizure. Three representative patients are shown (right: S01, matched PUL, unmatched ANT; middle: S22, matched ANT, unmatched PUL; left: S12, matched PUL, unmatched VIM/VOP). (**c**) Raw traces of h2 correlation coefficient (smoothed for visualization) for a representative seizure for each patient of panel b. The gray shaded area includes the seizure duration (from onset to termination). (**d**) Schematic illustrating the hodologically matching (bold colored lines) and unmatching (dashed gray lines) thalamocortical connections. (**e**) Violin plot showing the mean h2 coefficient across seizures for each matched (top) and unmatched (bottom) thalamus-SOZ pair. (**f**) Box plot of mean h2 coefficient across seizures during initiation, middle and termination phase of each seizure. (**g**) Percentage difference of the change in h2 coefficient during seizures between matched and unmatched nuclei, across different frequency bands of interest. For all boxplots, the whiskers extend to the maximum spread not considering outliers, central, top, and bottom lines represent median, 25^th^, and 75^th^ percentile, respectively. For all panels, statistical significance was assessed with two-tail bootstrapping with Bonferroni correction: p<0.05 (*), p<0.01 (**), p<0.001(***).

To computer the change in h2 between matched and unmatched nuclei (**Figure 3g**), we calculated, for each matched and unmatched pair, the percentage increase of h2 in the seizure duration with respect to the baseline epoch, as:

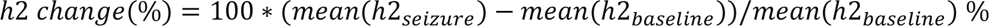

We then performed the subtraction between the percentage h2 changes calculated in matched thalamus-SOZ and unmatched thalamus-SOZ pairs.

For the correlation between the thalamus and non-SOZ regions, we randomly selected two seizures for each patient. We computed the mean h2 of the thalamus with 3 distinct non-SOZ pair of contacts (**Supplementary Table 3**).

#### Granger Causality analysis

To analyze the directionality of the interactions between the thalamus and the cortex, we used the Granger Causality method^39^. First, we selected all the seizures that lasted at least 10 s and showed an increase in the h2 correlation coefficient in the theta to beta band. We filtered the raw SEEG signals in the 4-30 Hz range and divided each seizure recording in 10 s non-overlapping windows. We computed the GC matrix of each matched thalamus-SOZ pair for every epoch. This computation results in a 4X4 matrix for each epoch, where the second diagonal contains the thalamocortical (th→SOZ) and corticothalamic (SOZ→th,) GC value. In addition to the absolute GC value, we quantified per percentage change in GC from initiation to termination (**Figure 4 c, d, f**) as:

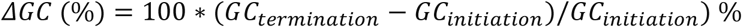

where termination and initiation are the 10 seconds epochs preceding the seizure onset and the seizure termination time stamps, respectively. To avoid misinterpretations given by the variability in the number of seizures recorded from each patient, we highlight the single-subject results in **Extended Data Figure 4**. In a total of 20 patients, 17 showed an increase in the proportion of seizures with the thalamus leading at termination (pie charts in **Extended Data Figure 4**) and 18 showed higher *ΔGC* in the thalamocortical (and not corticothalamic) coefficient (bar plot in **Extended Data Figure 4**). Such finding robustly confirms population-observed trends.

**Figure 4:**
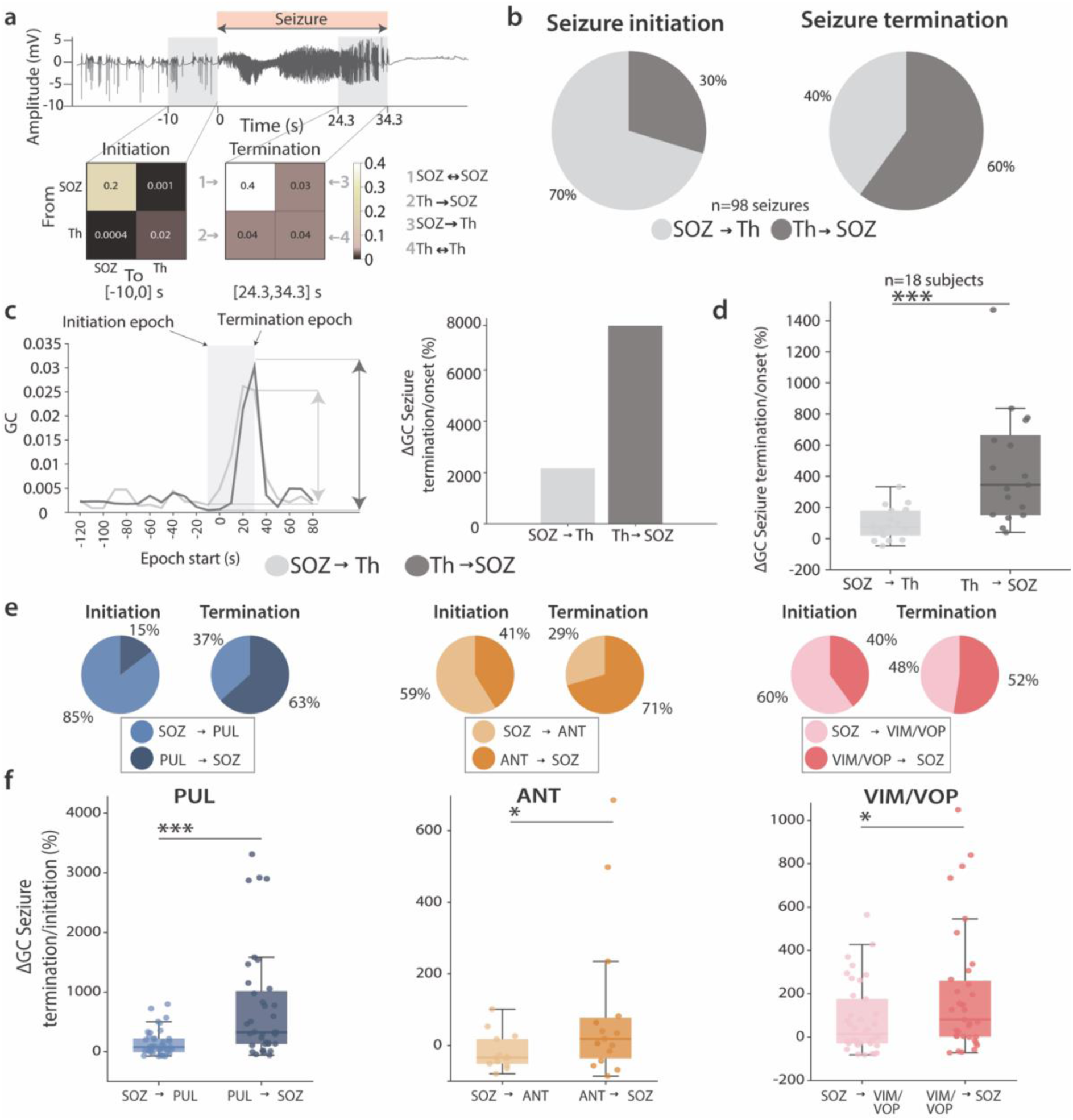
Analysis of thalamocortical versus corticothalamic interactions during seizures. (**a**) Representative seizure recording (S01). The red bar indicates the full seizure duration, while the shaded gray regions show the initiation and termination epoch. On such windows, we calculated the GC coefficient matrix that illustrates connectivity from and to the thalamic nuclei to a matched SOZ. (**b**) Pie charts illustrating the proportion of seizures with higher corticothalamic (SOZ→th, light gray) or thalamocortical (th→SOZ, dark gray) GC coefficient for the initiation (left) and termination (right) epoch. (**c**) (*Left*) Plot of the evolution of corticothalamic and thalamocortical GC coefficients for 10 s epochs starting two minutes before a representative seizure. The arrows indicate the relative change of GC from initiation to termination. (*Right*) Quantification of the relative percentage change of GC coefficient from initiation to termination, for corticothalamic (light gray) and thalamocortical (dark gray) direction. (**d**) Boxplot of the relative percentage change of GC coefficient from initiation to termination for all subjects. Each data point represents the mean value across all the seizures for each subject. (**e**) Pie charts illustrating the proportion of seizures with higher corticothalamic (SOZ→th, light color) or thalamocortical (th→SOZ, dark color) GC coefficient for the initiation (left) and termination (right) epoch, separated from the three nuclei of interest (PUL at left, ANT at middle, VIM/VOP at right). (**f**) Boxplot of the relative percentage change of GC coefficient from initiation to termination for all the seizures separated for the matched thalamic nucleus investigated (PUL at left, ANT at middle, VIM/VOP at right). For PUL n=38 and n=36 seizures, for ANT n=14 and n=15 seizures, for VIM/VOP n=38 and n=35 seizures. For all boxplots, the whiskers extend to the maximum spread not considering outliers, central, top, and bottom lines represent median, 25th, and 75th percentile, respectively. Outliers were removed with quartile method (see Methods). For all panels, statistical significance was assessed with two-tail bootstrapping with Bonferroni correction: p<0.05 (*), p<0.01 (**), p<0.001(***).

### Case report: CM connectivity in generalized epilepsy

We had the unique opportunity to justify the exclusion of the CM in our study (**Extended Data Figure 1**), focused on focal epilepsy, by performing structural and functional connectivity analysis of the CM, as well as the PUL, ANT and VIM/VOP on a patient affected by generalized epilepsy. In particular, the patient was a female in her late 20s, with a severe medically refractory epilepsy, with more than 10 generalized seizures per day. The patient underwent SEEG exploration to investigate the appropriate thalamic target for neuromodulation as a part of her clinical care.

We performed h2 correlation analysis of 24 seizures recorded with simultaneous sampling of CM, PUL, ANT, VIM/VOP as well as 13 bipolar electrodes sampling the following brain structures: insula, frontal operculum, superior temporal sulcus, superior temporal gyrus, middle temporal gyrus. We computed the mean h2 coefficient for each thalamic nucleus with the cortical electrodes. Unlike the focal epilepsy patients, where the SOZ only comprises a pair of cortical electrodes, here we averaged the h2 values of each thalamus-cortex pair.

### High-frequency thalamic stimulation

#### Stimulation testing and protocol

During post-operative monitoring in the EMU, we had the unique opportunity to perform acute thalamic stimulation testing on 14 patients of our cohort. Importantly, we could stimulate in both matched and unmatched thalamic nuclei. We applied continuous electrical stimulation with bipolar configuration on the thalamic site of interest, at a frequency of 100 Hz. This value was chosen because it was the highest allowed by the stimulator used, and it is in line with commonly used parameters for thalamic electrical stimulation (≥100Hz). The amplitude and pulse width were adjusted individually for each patient according to their comfort levels, but never exceeded the ranges of 1-3 mA and 60-300 us respectively.

For clinical safety reasons, the stimulation testing occurred only after the patients exhibited spontaneous seizures and in seizure-free days. The duration of the stimulation train varied from a minimum of 5 seconds to a maximum of 30 seconds, repeated over 1-10 times depending on the patient comfort and clinical state.

#### Preprocessing and IED detection

To analyze the immediate effect of thalamic stimulation we extracted the following data: i) for baseline (Stim OFF), we collected 2 minutes of interictal activity in the SOZ channels before any kind of stimulation was delivered to the patient; ii) for active stimulation period (Stim ON), we included the SOZ recording for the whole duration of stimulation up to 10 s after stimulation was stopped^33^. We excluded the first two seconds after the stimulation was delivered to reduce false positive detection due to transient stimulation-induced burst^40^. Unfortunately, for technical reasons, 4 patients had a shorter baseline period (average of 45 s). Prior epoching, we preprocessed the data in Anywave^38^ by applying a band-pass filter (1-1000 Hz) and a notch filter (60Hz) and manually removing channels affected by stimulation artifact or other noise sources. On the remaining channels, we applied independent component analysis (ICA) and extracted n components (where n=number of channels −1). This method is a common technique used to separate independent sources linearly mixed in several sensors. We visually inspected all the components and rejected the ones associated with stimulation artifacts. In this way, we could reliably apply an automatic spikes detection algorithm on the stim ON and stim OFF recordings.

Each intracranial recording of interest was then divided in 5 s non-overlapping epochs, on which we computed the detection of epileptiform discharges. To avoid any confound due to the stimulation artifact, we considered as IED only interictal spikes, and not high-frequency oscillations. For this, we used Delphos software^41,42^, which detects interictal spikes from SEEG recording from their time-frequency representation. This detector was previously demonstrated to allow for almost 100% specificity and more than 80% sensitivity. To ensure correct spikes detection we 1) computed the analysis also on non-SOZ channels, to confirm that no spikes would be detected and 2) visually inspected the data. We saved the IED number for each Stim ON and Stim OFF epoch.

#### IED analysis

To quantify the impact of acute thalamic stimulation on the IED rate, we computed two complementary analyses, based on the data quality: a) if the IED were clearly detectable and frequent (occurring in at least 50% of the baseline epochs), we calculated the IED rate as the number of IED per minute, in each Stim ON and Stim OFF epoch; b) if more than half of the baseline epochs presented no IED, we calculated the probability of finding no spikes across all the Stim ON and Stim OFF epochs (**Supplementary Table 2**). Even if consistent spiking was observed in baseline epochs, we included in analysis b also a subject (S26) that presented complete suppression of IEDs in stimulation epochs, hence precluding statistical analysis. For both analyses, we computed the percentage difference of these quantities for Stim ON vs stim OFF to characterize the stimulation effect. A beneficial effect of stimulation on epileptiform discharges would result in a smaller IED rate with stimulation ON for analysis a, and a positive change in analysis b, prompting to show that stimulation epochs would more likely show complete suppression of IED.

### Clinical outcome of chronic thalamic stimulation

We evaluated clinical outcome of hodologically matched PUL and ANT stimulation in seven individuals (S16, S27-S32) who received a chronic neuromodulation system as part of their clinical care.

Demographics and epilepsy information for these patients is reported in **Supplementary Table 1**. All patients received a bilateral implant of the PUL or ANT, according to their SOZ location and as indicated by their clinical team following SEEG monitoring. Six patients received a Responsive Neurostimulator (RNS), while one patient received a DBS system (Medtronic Percept). Stimulation parameters were individually tailored for each patient according to their clinical need (**Supplementary Table 4**). No clinical decisions were based upon this research.

We retrospectively evaluated the percentage reduction in disabling seizures’ frequency and the change in antiseizure medication. The follow-up window ranged from 9 to 30 months.

### Data analysis and statistical procedures

All figures and analysis were performed using Matlab 2023b (Mathworks, California, US). For all box plots reported in this manuscript, the whiskers extend to the maximum spread not considering outliers, central, top, and bottom lines represent median, 25^th^, and 75^th^ percentile, respectively.

For all the analysis, we used the bootstrap method, which does not rely on distributional assumptions of the data, but rather resamples the quantities of interest to achieve empirical confidence intervals. For each comparison, we created a bootstrap sample (for n=10000 repetitions) by drawing a sample with replacement from the actual data points (n=10000 repetitions), and calculated the difference in means of the resampled data. We then applied two-tailed bootstrapping with significance levels of 0.05 (95% confidence interval), 0.01 (99% confidence interval), or 0.001 (99.9% confidence interval). The null hypothesis of no difference in the mean was rejected if 0 was not included in the confidence interval of the corresponding alpha value. If multiple comparisons were performed at once, we used a Bonferroni correction by dividing the alpha value by the number of pairwise comparisons being performed.

## Results

### Study rationale and design

We aimed at investigating whether specific thalamic subnuclei might provide increased efficacy in the treatment of focal refractory epilepsy based on their hodological matching with the cortical SOZ. To demonstrate our hypothesis, we deployed a large array of electrophysiological and imaging assessments in a unique cohort of 32 focal MRE patients (**Figure 1a** and **Supplementary Table 1** for demographic and clinical information for each patient). Specifically, we analyzed intracranial electrophysiological data from 26 patients (S01-S26) undergoing (SEEG) exploration for epilepsy monitoring (**Figure 1b**). Prior to SEEG implantation, we acquired preoperative structural magnetic resonance imaging (MRI) and postoperative computed tomography (CT) for precise SEEG contacts localization performed by a certified neurosurgeon (JGM) in all patients. All the participants had at least one SEEG electrode located in one of the nuclei of interest, namely the PUL, ANT or VIM/VOP nuclei, and a diagnosis of focal epilepsy. Overall, 37 thalamic nuclei among those of interest were sampled in this cohort (**Figure 1b, Supplementary Table 2**). We categorized each patient’s SOZ as frontal (meaning originating from frontal lobe regions such as the orbitofrontal cortex), temporal (including mesial and lateral temporal lobe structures), rolandic (including pre-and post-central peri-rolandic regions) or parietal/occipital cortex (namely originating from posterior-quadrant areas) (**Figure 1c, Supplementary Table 3**). In a subgroup of patients (n=8), we acquired and analyzed high-resolution diffusion MRI data to corroborate the anatomical organization of thalamocortical fibers (**Figure 1f**). Such connectivity pattern was also confirmed through electrophysiology with thalamocortical evoked potentials (n=6). To study the interaction of the thalamus and the epileptic cortex, we then analyzed a total of 216 spontaneous seizures with concurrent sampling of the thalamic nuclei and cortical SOZ. Hence, each recorded seizure was categorized according to the SOZ anatomical location and the sampled thalamic nucleus during the event (**Figure 1c**). We then performed electrophysiological testing in a subset of patients. Specifically, we used high-frequency thalamic stimulation (n=14) to test immediate effects of hodologically-matched thalamic stimulation on interictal epileptiform activity (**Figure 1f**). Finally, we assessed long-term efficacy of our hodology based neuromodulation approach in 7 patients (S16, S27-32) with a chronic stimulation implant (**Figure 1d**) and up to 3 years of follow-up (**Figure 1e**).

Importantly, we also reported a single case of generalized epilepsy with simultaneous sampling of PUL, ANT, VIM/VOP as well as CM: this last site showed widespread structural and functional connectivity (**Extended Data Figure 1)**, replicating well known evidence from the literature^8,11,22,43^ and supporting its applicability as a target in generalized, rather than focal, epilepsy.

Overall, we designed and performed a comprehensive study to investigate the mechanisms and efficacy of hodologically-matched thalamic stimulation in focal refractory epilepsy.

### A specific hodological map of the thalamocortical pathway in the epileptic brain

While previous studies proposed a structural connectivity map of the thalamocortical projections ^17,44,45^, the majority of these studies were performed in healthy brains and were limited to imaging techniques, such as fiber tracking. Here, we confirmed the presence of a well-defined anatomical map of the thalamocortical projections of PUL, ANT and VIM/VOP in the epileptic brain with two complementary methodologies: neuroimaging and electrophysiology.

First, we performed high-definition fiber tracking (HDFT) of high-resolution diffusion MRI data to confirm previously reported anatomical organization of cortical connectivity patterns of the PUL, ANT and VIM/VOP. We reconstructed all the likely axonal pathways between these nuclei and the cortical regions presenting seizures in our cohort (namely frontal, rolandic, parietal, occipital and temporal) (**Figure 2a**). We quantified the relative strength of these connections by calculating the volume of thalamocortical projections from each nucleus to each cortical region normalized by the total volume of fibers (**Figure 2b**). This analysis revealed a clear thalamocortical anatomical organization. Indeed, the PUL showed preferential connectivity towards the parietal and occipital regions, and the temporal lobe (mean volume value, frontal 0.11, rolandic: 0.04, *parietal*: 0.38, *occipital*: 0.27, *temporal*: 0.18); the ANT, instead, presented axonal projections going preferentially towards the frontal and temporal lobes (*frontal: 0.39*, rolandic: 0.05, parietal: 0.06, occipital: 0.16, *temporal: 0.31*); finally the VIM/VOP nuclei projected more remarkably to the rolandic cortex (frontal: 0.29, *rolandic: 0.56*, parietal: 0.03, occipital: 0.06, temporal: 0.03). This constricted structure of thalamocortical fibers from PUL, ANT and VIM/VOP contrasts with the broad anatomical connectivity of the CM (frontal: 0.27, rolandic: 0.17, parietal: 0.19, occipital: 0.19, temporal: 0.15), hence corroborating its indication as a more suitable target in generalized epilepsy (**Extended Data Figure 1a-b**).

We then confirmed this anatomical organization by analyzing thalamocortical evoked potentials (EPs) in the different cortical areas. For this, we applied low frequency (1 Hz) electrical stimulation to the different thalamic nuclei at 1-3 mA while recording cortical neural response evoked by each pulse in the different cortical lobes (**Figure 2 d, g, j**). We then analyzed stimulation triggered averages of EPs. Following thalamic stimulation, we observed robust cortical EPs in anatomically connected (i.e., matching), but not in unmatching, cortical regions (**Figure 2 e, h, k**). Indeed, analysis of EPs peak to peak amplitudes showed significantly larger responses in those areas that matched the previously identified projections, further confirming a precise anatomical organization of distinct thalamic nuclei (**Figure 2 c, f, i, l**).

In summary, we confirmed with multiple techniques a specific anatomical organization of the thalamocortical projections of PUL, ANT and VIM/VOP in the studied population. We then define hodologically-matching (parietal, occipital and temporal for PUL, frontal and temporal for ANT, and rolandic for VIM/VOP) and unmatching (frontal and rolandic for PUL, rolandic, parietal and occipital for ANT, frontal, temporal, parietal and occipital for VIM/VOP) regions.

### Increased thalamocortical coupling during seizures in hodologically-matching SOZs

Given this specific structural organization of the thalamocortical fibers, we hypothesized a preferential functional involvement in the course of spontaneous seizures of the thalamic subnucleus that anatomically matches the SOZ (**Figure 3d**). To test this hypothesis, we analyzed a total of 216 spontaneous (i.e., not induced by cortical electrical stimulation) ictal events. The anatomical location of the SOZ as well as the relevant time markers (pre-ictal, beginning and end) of each seizure were determined by a certified clinical team. For each seizure we computed the well validated non-linear h2 correlation coefficient^29,46–48^ to identify time-resolved synchrony between each thalamic nucleus and each cortical SOZ. Specifically, we computed mean h2 coefficient during the ictal event (from seizure onset to termination) and the mean h2 coefficient during a 10 second baseline epoch spaced 2 minutes apart from the seizure’s onset^29^. We then compared mean h2 coefficient between the two phases separately for anatomically connected and non-connected thalamic nuclei to quantify the degree of specificity of thalamic involvement during spontaneous seizures.

As hypothesized, we found that while anatomically matched nuclei showed a significant increase of synchrony between the thalamus and the SOZ, indicating functionally specific thalamic involvement during ictal events (**Figure a**), minimal or no correlation was found in unmatched nuclei (**Figure 3 b, c**). This was consistent for all thalamic nuclei of interest and subjects (**Figure 3e**, mean h2 was 0.13 vs 0.20 in matched with p<0.001, and 0.12 vs 0.11 with p>0.05 in unmatched, for baseline and seizure respectively). Importantly, we also observed that thalamic nuclei had minimal or no correlation with non-SOZ regions (**Extended Data Figure 3 a, b**), demonstrating the specificity of the correlation with the anatomically-matched SOZ. Additionally, for the patient diagnosed with generalized epilepsy we found that PUL, ANT, and VIM/VOP nuclei had a lower correlation with the epileptogenic zone (0.16, 0.25, 0.21 mean h2 across 24 seizures and 13 channels for PUL, ANT, and VIM/VOP, respectively) as compared to the CM (0.3, **Extended Data Figure 1 c, d**). This confirms the more diffused profile of the CM connectivity (both anatomically and functionally), entailing its suitability for generalized, rather than focal epilepsy treatment. Overall, these functional outcomes robustly validated the thalamocortical anatomical organization observed with fiber tracking, further supporting the proposed definition of hodologically-matching and unmatching nuclei.

To further investigate possible mechanisms of thalamocortical interactions during focal seizures, we explored the coupling of the thalamus and SOZ in terms of seizure phases and frequency bands for matched thalamic nuclei. In particular, we aimed at verifying that the mechanism of thalamic involvement during seizures would not differ across the three nuclei of interest, since a different mechanism could imply a different design of electrical stimulation therapies. First, we asked whether maximal coupling of matched nuclei was specific to distinct seizure phases. To this aim, we computed the mean h2 correlation coefficient in three epochs: seizure initiation (including the 10 seconds preceding the first appearance of a tonic discharge in the SOZ contacts), middle part of the seizure (defined as the time interval separating seizure initiation and termination), and seizure termination (which includes the last 10 seconds of discharge). We found that the highest synchrony between the thalamus and SOZ occurred at seizure termination (**Figure 3f**). Importantly, this behavior was present for all three thalamic nuclei when hodologically (i.e., anatomically and functionally) matched to the SOZ (**Extended Data Figure 3c**). These findings support and validate the notion that synchronization loops between the thalamus and cortex might contribute to the termination of seizures, as suggested in previous studies in animals^29,49^ and humans^50,51^.

Additionally, we sought to determine if the observed synchrony presented a specific spectral profile. Hence, we compared the mean h2 coefficient computed in different frequency bands of the intracerebral recordings (delta, theta, alpha, beta, low gamma, high gamma, ripple, fast ripple). No spectral specificity was revealed: indeed, for all the frequency bands except the fast ripple band, the correlation between hodologically-matched thalamic nuclei and SOZ always increased significantly during the ictal event, while it showed no variation for unmatched thalamic nuclei (**Extended Data Figure 3d**). Hence, we asked whether specific frequency bands would better distinguish hodologically-matched and unmatched nuclei potentially suggesting biomarkers for the refinement of the stimulation design. For this, we compared the increase in ictal correlation respect to baseline between hodologically-matched and unmatched nuclei in different frequency bands. Interestingly, we found that theta, alpha and beta frequencies had a higher difference between the h2 correlation change during seizures between hodologically-matched and unmatched nuclei (**Figure 3g**). These results show that the theta-beta frequencies might be more relevant to differentiate thalamic nuclei based on their connectivity to the SOZ.

All in all, these findings demonstrate that thalamic nuclei exhibit enhanced neural synchrony with hodologically-matched cortical SOZs, particularly at the period of seizure termination, and primarily driven by theta, alpha, and beta oscillations.

### Thalamocortical synchrony revers directionality during spontaneous seizures

While the h2 index revealed an increase in correlation between the hodologically-matched thalamic nuclei and the SOZ throughout the seizure course, here we sought to determine the directionality of this coupling. Importantly, previous studies reported a change in the directional coupling of thalamocortical interactions in temporal lobe epilepsy throughout the seizure course^29,52^. Specifically, thalamic involvement was shown to occur early in a seizure, although it did not typically lead at the onset^29^. Additionally, animal studies have shown that the thalamus participates during seizures and plays a crucial role in their development, but it is less involved at seizure onset^49^. In order to explore whether a similar behavior was prevalent for all thalamic structures here investigated, we further expanded the analysis of hodologically-matched thalamocortical synchrony during spontaneous seizures by investigating the directionality of the coupling. To this purpose, we used Granger Causality (GC)^39^ between the thalamic nuclei of PUL, ANT and VOP/VIM and the hodologically-matching SOZs. This methodology could indeed provide insights in the understanding of propagation patterns of epileptic activity helping the future design of stimulation patterns aimed at decreasing epileptiform activity. Since our previous results suggest a crucial role in the theta-alpha-beta oscillations, for this analysis we included only band-passed filtered (4-30 Hz) seizures that showed a clear increase in h2 coefficient in this frequency range and lasted at least 10 seconds. This resulted in a total of 98 seizures. For each seizure, we extracted two 10 s epochs of interest at seizure initiation and seizure termination and computed the GC matrix for the SOZ contacts and the hodologically-matching thalamic nucleus contacts (**Figure 4a**). Importantly, GC of unmatched nuclei was not explored because of the lack of correlation with SOZ. To test whether the role of different thalamic nuclei was changing over the course of the seizure, we compared the GC coefficient from the thalamus towards the SOZ (th→SOZ) and from the SOZ towards the thalamus (SOZ→th).

At seizure initiation, in the majority of the seizures (70%), the GC coefficient of SOZ→th was higher than the GC coefficients th→SOZ (**Figure 4b** for all subjects together**, Extended Data Figure 4** for each individual) suggesting a leading role of the cortex. In contrast, more than half of the events (60%) presented a higher th→SOZ coefficient at the termination of ictal events hence indicating a change in the leading anatomical structure. This could be due to 1) SOZ→th decreasing while th→SOZ remaining stable (or decreasing less) throughout the course of the seizure, 2) th→SOZ increasing and SOZ→th remaining stable (or increasing less). To test for this, we computed the relative change of the GC coefficient between termination and initiation and compared it between SOZ→th and th→SOZ. Interestingly, we noticed an increase in both the thalamocortical (th→SOZ) and corticothalamic (SOZ→th) coupling throughout the duration of the event (**Figure 4c**). However, the relative change was higher for the th→SOZ consistently across all the subjects, further demonstrating a leading role of the thalamus at seizure termination (73.41 vs 345.6%, for SOZ→th and th→SOZ, respectively) (**Figure 4 c, d**).

Importantly, we verified that the same characteristics of thalamocortical interactions were not specific to a particular nucleus. As a matter of fact, in all studied nuclei, the absolute GC value in the initiation epoch and termination epoch showed a reverse trend, with a primary leading role of the cortex at first (85% for PUL, 59% for ANT, 60% for VIM/VOP), followed by a predominantly leading role of the thalamus at termination (63% for PUL, 71% for ANT, 52% for VIM/VOP) (**Figure 4e**). Additionally, PUL, ANT and VIM/VOP showed a significantly higher relative change for th→SOZ throughout the seizure (**Figure 4f**).

In summary, here we investigated the underlying mechanisms of thalamocortical and corticothalamic coupling involving the ANT, PUL and VIM/VOP with hodologically-matched SOZ during spontaneous seizures. Our findings suggest a potential trend where cortical regions exert a dominant influence at seizure initiation, while the thalamus may assume a more prominent role as the ictal events progresses. This pattern was observed across all three nuclei, suggesting that the development of nuclei-specific stimulation protocols-particularly regarding the timing of stimulation during the ictal event-may not be necessary to address the varying underlying mechanisms of thalamic action.

### Matched thalamic stimulation immediately suppresses epileptiform discharges

Given the peculiar hodology that we previously found both in the structural connectivity of the PUL, ANT and VIM/VOP, as well as in the functional coupling observed during ictal events, we hypothesized that targeting electrical stimulation to the thalamic nucleus that hodologically-matches the cortical SOZ might be more effective in reducing epileptiform activity than unmatched nuclei. To test this hypothesis, we delivered bipolar thalamic stimulation through the SEEG electrodes in 14 patients. For clinical reasons, the stimulation testing was always performed after the patient had shown spontaneous seizures. We evaluated the effect of hodologically-matched and unmatched stimulation on interictal epileptiform discharges (IEDs), namely interictal spikes. We used an automatic validated algorithm^41,42^ to extract IEDs from cortical recordings in the SOZ (**Figure 5a**), previously subdivided in 5 seconds epochs and denoised (see **Methods**). We aimed at quantifying the difference between baseline (including the epochs in the 2 minutes preceding the first stimulation applied) and stimulation epochs (see **Methods**). For this, we performed two distinct analysis: a) in n=8 patients with continuous evident pathological spiking (**Figure 5a**), we computed the rate of IEDs per minute in baseline (Stim OFF) and stimulation (Stim ON) epochs; b) in n=6 patients with more than half of the baseline epochs without pathological spikes, we computed the probability of suppressed IEDs in stimulation epochs and compared it to the probability of suppressed IEDs in baseline epochs, hence obtaining a percentage change in IED suppression probability (**Supplementary Table 2**). In the first analysis, a positive effect of the stimulation (meaning reducing pathological IEDs) should result in a smaller IEDs rate during stimulation epochs than during baseline (stimulation off) epochs; whereas in the latter, a positive number indicated that the stimulation epochs are more likely to present no IEDs than the baseline epochs (without stimulation).

**Figure 5:**
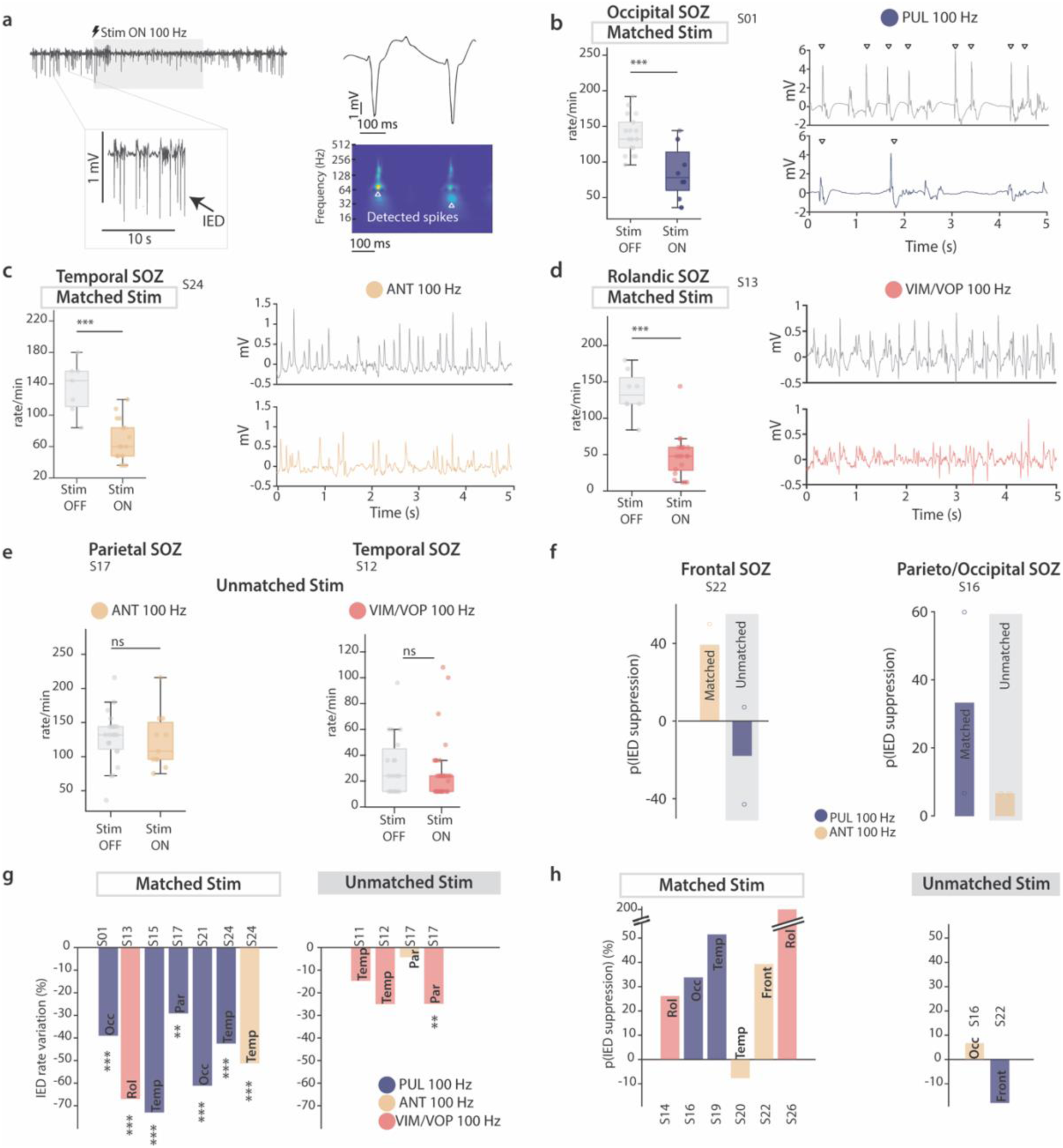
Hodologically-matched electrical stimulation of the thalamus better reduces pathological IEDs. **(a**) Representative example of an interictal SEEG recording of a SOZ contact, with clearly visible IED (spikes) and their time-frequency representation exploited for automatic detection. (**b**) (*Left*) boxplot for IED rate/minute in stimulation OFF (n=18) and stimulation ON (n=8) epochs of the PUL (matched nucleus). (*Right*) representative stim OFF (*top*) and stim ON epoch (*bottom*) with matched PUL stimulation shows immediate suppression of IED. (**c**) Representative example, as in panel b, for matched ANT stimulation (n=7 and n=17 for stimulation OFF and ON, respectively). (**d**) Representative example, as in panel b and c, for matched VIM/VOP stimulation (n=8 and n=21 for stimulation OFF and ON, respectively). (**e**) Boxplot of representative examples of unmatched thalamic stimulation on IED rate for ANT (left, n=18 vs n=8 for stimulation OFF and ON, respectively), and VIM/VOP (right, n=19 vs n=41 for stimulation OFF and ON, respectively). (**f**) Percentage change relative to baseline of the probability of IEDs suppression in matched and unmatched thalamic stimulation in two representative patients. (**g**) All-subject analysis of the IED rate percentage variation to baseline with matched (*left*) and unmatched (*right*) stimulation. (**h**) All-subject analysis of the percentage change relative to baseline of the probability of IEDs suppression in matched (*left*) and unmatched (*right*) stimulation. For all boxplots, the whiskers extend to the maximum spread not considering outliers, central, top, and bottom lines represent median, 25th, and 75th percentile, respectively. Outliers were removed with quartile method (see Methods). For all panels, statistical significance was assessed with two-tail bootstrapping with Bonferroni correction: p<0.05 (*), p<0.01 (**), p<0.001(***).

As hypothesized, we found that electrical stimulation of the PUL, ANT and VIM/VOP significantly suppressed cortical IEDs when the SOZ was in a cortical region that hodologically-matched the thalamic nucleus stimulated (**Figure 5 b-d, g**). Overall, our analysis revealed that hodologically-matched but not unmatched thalamic stimulation significantly reduced the IED rate in all tested patients (for matched stimulation, IED variation to baseline ranged from −29.12% to −73% with p<0.01 and p<0.001, while for unmatched stimulation, it ranged from −4.2% to −25%) (**Figure 5g**). Here, we highlight the IED rate values for a representative patient for each nucleus of interest: we observed a reduction of the IED rate of 132 vs 78 spikes/minute for the PUL (median rate at stimulation off vs stimulation on in S01, p<0.001) (**Figure 5b**), 144 vs 60 spikes/minute for the ANT (in S24, p<0.001) (**Figure 5c**), and 132 vs 48 spikes/minute in the VIM/VOP (in S13, p<0.001) (**Figure 5d**). Similarly, when evaluating the percentage change in the probability of IED suppression, we found that hodologically-matched thalamic stimulation epochs were more likely to present no IEDs when compared to unmatched thalamic stimulation epochs. Indeed, In 5 out of 6 patients that received stimulation in the nucleus matching their SOZ, we found at least a 20% increase (up to over 200%) in the probability of epochs with no IEDs with stimulation on with respect to stimulation off (**Figure 5h**); whereas the stimulation effect was more inconsistent (−17 to +7%) when the targeted nucleus was not matching the hodology of the thalamocortical fibers towards the SOZ (**Figure 5h**). This finding further suggests that the more effective thalamic target to suppress IEDs is the one matching the hodology of the SOZ.

Overall, we showed that only targeted electrical stimulation of the nuclei hodologically-matching the SOZ is significantly reducing the rate of IEDs and increasing the probability that stimulation epochs present no IEDs (complete suppression). These findings are crucial in guiding approaches to address the heterogeneity of SOZs and in clinically selecting optimal stimulation targets for the electrical neuromodulation of focal epilepsies.

### Matched thalamic electrical stimulation in chronic epilepsy treatment

Finally, to verify whether the observed immediate effects on IEDs could be translated in a long-term clinical improvement in epileptic patients, we evaluated 7 patients (one of which being part of the SEEG cohort) that were chronically implanted for thalamic stimulation in a hodologically-matching thalamic nucleus, as part of their clinical care. Across all patients, 6 (S16, S27-S31) presented focal MRE originating from the posterior quadrant regions and hence received a neurostimulation system in the PUL, while one (S32) was implanted in the ANT to treat a frontal onset MRE (**Supplementary Table 4**). Overall, we observed a median reduction in seizure frequency of 86.5 % (**Figure 6a, b**) at 15.86±7.9 (mean±SD) months of follow-up. As a representative example, for S16 seizures reduced from 5 per week to 1 per month, and the patient also self-reported a significant reduction in seizure severity, noting no missed workdays since the implant of the stimulation device, compared to missing 1-2 days per month prior to surgery. Additionally, the anti-seizure medication (ASM) intake for each patient decreased for 5/7 patients since the matched neurostimulation system was implanted, and did not increase for any of the participants (**Figure 6c**). Importantly, these results dramatically outperform the unmatched thalamic stimulation results reported in literature for the SANTE trial at 7 years follow-up. Indeed, in this previous study, unmatched stimulation (ANT stimulation for seizure onsets other than frontal or temporal) only achieved 39% reduction in seizure frequency, whereas outcome for matched SOZ were comparable to the ones we observed (78% and 86% for temporal and frontal onset, respectively)^26^.

**Figure 6:**
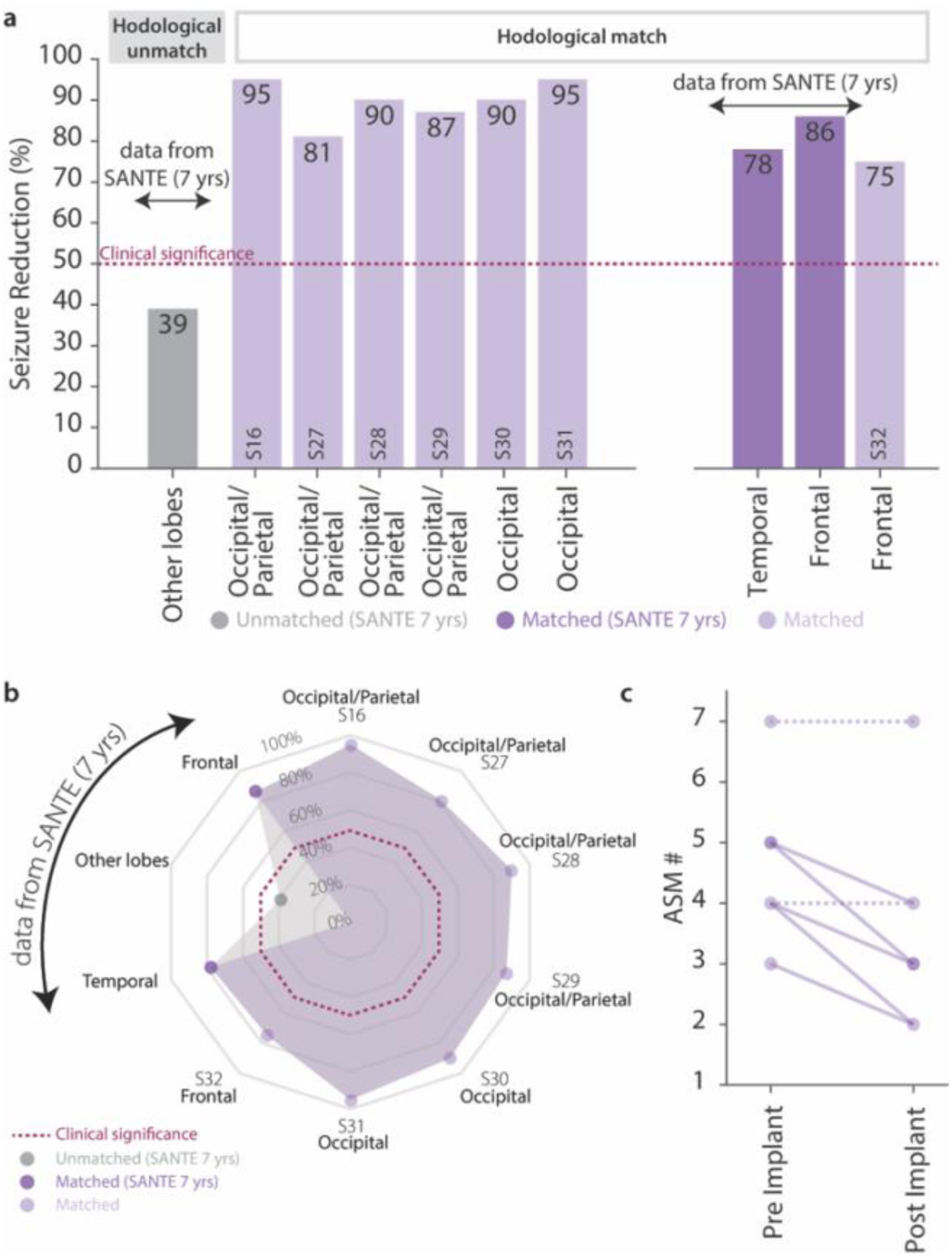
Hodologically-matched thalamic stimulation as a neuromodulation therapy in chronic epilepsy treatment. (**a-b**) Bar plot and radar plot of the percentage of seizure frequency reduction for S16, S27-S32, compared to the median seizure frequency reduction reported by the SANTE trial at 7 years follow up for frontal, temporal and other lobes. (**c**) Scatter plot of the number of antiseizure medication for each patient prior and after the matched thalamic neurostimulation implant. Dashed lines indicate no change in the numbers of ASM.

In conclusion, our results indicate that hodologically-matched thalamic stimulation provides significant therapeutic benefits, including notable reductions in the frequency and severity of disabling seizures. This approach not only outperforms current standard treatments but also lays the groundwork for more personalized and precise target selection in neuromodulation for medically refractory focal epilepsy, particularly in patients who are not candidates for curative resective or ablative interventions.

## Discussion

Electrical stimulation of the thalamus is undergoing a rapid advance as a treatment opportunity for patients with MRE^12,53,54^, with indications in both generalized^22,53,55^ and focal epilepsy^9,56–60^. While in generalized epilepsy the CM nucleus effectively reduce seizure events, in focal epilepsy, this expansion requires a profound understanding of the intricate anatomical and functional thalamocortical interactions with highly heterogeneous and individualized epilepsy phenotypes to improve clinical outcome. Here, we proposed a novel approach to select the optimal targets for thalamic electrical stimulation in focal epilepsy, considering the specificity of SOZs in relation to cortical and subcortical anatomy. Specifically, we demonstrated that leveraging the hodology-based interactions between certain subnuclei of the thalamus and the epileptogenic cortex results in stronger neuromodulator efficacy. This approach highlights the importance of thalamocortical hodology in individualizing stimulation targets, potentially advancing therapeutic outcomes, and paving the way for more effective neuromodulation strategies in patients with focal MRE who are not candidates for curative interventions.

Since the early stages of epilepsy surgery and the first mechanistic studies about epileptogenicity, focus has increasingly turned towards deeper brain structures, particularly the thalamus^61,62^. Early animal studies by Penfield and Jasper in 1947 identified the thalamus as a potential generator of epileptic cortical activity, proposing the theory of “centrencephalic” epilepsy^18,63^. Indeed, this structure mediates reciprocal cortical-subcortical connections, functioning as an ‘integrative hub’ for functional brain networks^64^. Recently (challenging previous theories), the thalamus has been implicated not in the generation, but in the propagation of generalized and focal onset, due to its role in the bi-hemispheric cortical spread of epileptiform activity^32,50,65^. In addition, more recent evidence indicates that the thalamus plays a critical role also in the network dynamics of focal-onset epilepsies^19,29,31^, but lacked a consistent and systematic analysis of the hodological patterns of propagation from different cortical and thalamic regions during ictal events. Our results contribute to this discussion. Indeed, we detected significant correlation with ictal activity in all recorded thalamic nuclei further suggesting the thalamus as a potential neuromodulation target to treat focal MRE. However, correlation between thalamic and cortical SOZ activation was specific to thalamocortical anatomical connection, as identified by high-density tractography as well as electrophysiological assessment, supporting the hypothesis that epileptic propagation patterns during seizures obey predictable anatomo-functional patterns and highlighting the need to carefully select thalamic nuclei for neuromodulation according to individual clinical and electrophysiological data.

We applied this mechanistic understanding of the thalamocortical interactions to inform hodology-based electrical stimulation aimed at suppressing epileptiform activity. Our results demonstrated that hodologically-matched thalamic stimulation immediately suppresses interictal epileptiform discharges. Additionally, this targeted stimulation markedly enhances the likelihood of achieving complete suppression of IEDs during stimulation epochs, suggesting robust potential for targeted thalamic stimulation as a therapeutic strategy in epilepsy neuromodulatory management. Immediate effects in n = 14 subjects, paralleled with the drastic clinical improvement in seizure frequency with chronic matched thalamic stimulation (up to 95%) in seven patients, provide promising evidence that a hodology-based stimulation would improve clinical outcome for patients with focal epilepsy. Importantly, the comparative results show a substantial improvement over existing literature on unmatched thalamic neuromodulation (86.5% versus 39% reduction in seizure frequency), highlighting the potential need to reconsider current clinical practices, where a single target is often applied to all seizure onset zones.

Mechanistically, our results identified a trend suggesting that the epileptic cortex is the dominant structure at the initiation of seizures, while the thalamus assumes a leading role throughout the course of ictal events, showing higher synchronization towards the middle and termination of the ictal events. Importantly, this pattern was consistent across all thalamic regions investigated. In this regard, in an animal model of limbic epilepsy induced by GABA antagonist (i.e., bicuculline), Aracri and colleagues demonstrated hippocampal activation first, progressing to hippocampus-thalamus synchronization, and then thalamic bursts leading to the termination of seizures^66^. Destexhe and colleagues also highlighted the role of cortically induced coherence of thalamic-generated oscillations^67^. They implicated temporal and spatial thalamocortical interactions, suggesting that, at the onset of seizures, thalamocortical cells are hyperpolarized due to the greater power of excitatory cortical projections to reticular neurons compared to thalamic relay neurons. Once the initial cortical fast phase is abolished, thalamic relay neurons projecting to the cortex depolarize synchronically, reinforcing the coherence of seizure activity arising from the cortex, and promoting the highly synchronous cortical spike phase observed during the middle and end of seizures. Our findings related to the directionality of epileptic activity through thalamocortical circuits align with this mechanistic theory, where information flows to the thalamus from the epileptic cortex at seizure initiation and back to cortical areas during the late seizure phase. This supports the hypothesis that the thalamus plays distinct roles in this dynamic process^68^. Furthermore, it suggests the possibility to design closed-loop stimulation paradigm to restrict the stimulation to precise moment of the seizure development with potentially stronger neuromodulatory effects. In this regard, our findings indicate that the theta to beta frequency range represents a key hallmark of thalamocortical connectivity^69^, which could serve as a biomarker for seizure detection and closed-loop monitoring.

The primary limitation of this study is that the clinical assessment of matched electrical stimulation as a chronic therapy was evaluated in just seven patients as an observational retrospective report. Hence, inhomogeneous evaluation of all thalamic nuclei and SOZ occurred. However, these results are paralleled to a thorough investigation of the mechanisms of thalamocortical coupling in focal epilepsy, which support our findings. Additionally, even if limited in number, our evaluation of the effect of matched stimulation in a chronic stage of the treatment results in a drastic clinical improvement well above other literature reports. Larger clinical studies are now necessary to confirm the efficacy of hodologically-matched stimulation in the reduction of seizure frequency and improvements in patients’ quality of life in chronic settings. Another study limitation includes sampling bias associated with explorations of the human brain using intracranial electrodes, where conclusions can only be drawn from areas where electrodes were implanted. Nevertheless, strict inclusion and exclusion criteria minimized this limitation by focusing on subjects with highly localized seizures, many of whom became seizure-free after restricted focal resections. Additionally, the applied parameters of stimulation were restricted to few parameters (100 Hz only, 1-3 mA, continuous stimulation), mainly because the focus of the current study was related to the thalamic targeting and not to the different paradigms of electrical stimulation. However, several studies previously identified the optimal stimulation parameters in the high frequency range (>100Hz)^70–72^. Future studies will need to better refine stimulation parameters and identify optimal stimulation frequencies.

In summary, understanding the thalamocortical hodological principles governing the organization of focal seizures and their potential clinical applications is invaluable. Personalizing therapies based on the connectivity of the seizure onset zone to thalamic sub-nuclei and individual seizure characteristics may lead to improved efficacy and more favorable outcomes in the neuromodulation field for epilepsy. Our work addresses the critical unmet clinical need to identify optimal neuromodulation target by demonstrating predictable patterns of connectivity in thalamocortical interactions and guiding optimal thalamic targeting based on location of ictogenesis. This hodology-based approach would impact the current clinical standard in which one target fits all the SOZ (ANT in focal epilepsy, CM in generalized epilepsy), and lay the foundations for highly individualized neuromodulation treatments.

## Data Availability

All data produced in the present study are available upon reasonable request to the authors

## Author contributions

EP and JGM conceived the study. EP and JGM secured funding. AD, JGM, and EP designed the experiments. AD analyzed the data, with help from SN for correlation analysis and JH for DTI analysis. TA and JGM implemented patient recruitment, eligibility and monitoring and coordinated study management. AD, TA and JGM collected the data. AG, SS and ER helped with data management. AD, EP, JGM and TA helped with data interpretation. AD, JH, EP and JGM, wrote the paper and all authors contributed to its editing. All authors interpreted the data, contributed to the writing of the submitted manuscript, had final responsibility for the decision to submit the manuscript for publication, and vouch for the accuracy and completeness of the data and for the fidelity of the trial to the protocol.

## Acknowledgements

We would like to thank all the Epilepsy Monitoring Unit staff and Neurosurgery Staff of the University of Pittsburgh Medical Center for their support. We wish to thank all the participants and their families, who agreed to participate in the study and for the great insights into the real needs of people living with epilepsy.

## References

1. Kwan, P. & Brodie, M. J. Definition of refractory epilepsy: defining the indefinable? Lancet Neurol. 9, 27–29 (2010).

2. Thijs, R. D., Surges, R., O’Brien, T. J. & Sander, J. W. Epilepsy in adults. Lancet Lond. Engl. 393, 689–701 (2019).

3. Gupta, S., Ryvlin, P., Faught, E., Tsong, W. & Kwan, P. Understanding the burden of focal epilepsy as a function of seizure frequency in the United States, Europe, and Brazil. Epilepsia Open 2, 199–213 (2017).

4. Gloor, P. & Fariello, R. G. Generalized epilepsy: some of its cellular mechanisms differ from those of focal epilepsy. Trends Neurosci. 11, 63–68 (1988).

5. Badawy, R. A. B., Curatolo, J. M., Newton, M., Berkovic, S. F. & Macdonell, R. A. L. Changes in cortical excitability differentiate generalized and focal epilepsy. Ann. Neurol. 61, 324–331 (2007).

6. French, J. A. Refractory Epilepsy: Clinical Overview. Epilepsia 48, 3–7 (2007).

7. Li, A. et al. Neural fragility as an EEG marker of the seizure onset zone. Nat. Neurosci. 24, 1465–1474 (2021).

8. Velasco, F. et al. Deep brain stimulation for treatment of the epilepsies: the centromedian thalamic target. in Operative Neuromodulation: Volume 2: Neural Networks Surgery (eds. Sakas, D. E. & Simpson, B. A.) 337–342 (Springer, Vienna, 2007). doi:10.1007/978-3-211-33081-4_38.

9. Fisher, R. et al. Electrical stimulation of the anterior nucleus of thalamus for treatment of refractory epilepsy. Epilepsia 51, 899–908 (2010).

10. Fisher, R. S. Deep brain stimulation of thalamus for epilepsy. Neurobiol. Dis. 179, 106045 (2023).

11. Fisher, R. S. & Velasco, A. L. Electrical brain stimulation for epilepsy. Nat. Rev. Neurol. 10, 261–270 (2014).

12. Ryvlin, P., Rheims, S., Hirsch, L. J., Sokolov, A. & Jehi, L. Neuromodulation in epilepsy: state-of-the-art approved therapies. Lancet Neurol. 20, 1038–1047 (2021).

13. Shine, J. M., Lewis, L. D., Garrett, D. D. & Hwang, K. The impact of the human thalamus on brain-wide information processing. Nat. Rev. Neurosci. 24, 416–430 (2023).

14. Steriade, M. Synchronized activities of coupled oscillators in the cerebral cortex and thalamus at different levels of vigilance. Cereb. Cortex 7, 583–604 (1997).

15. Steriade, M. Sleep, epilepsy and thalamic reticular inhibitory neurons. Trends Neurosci. 28, 317–324 (2005).

16. Steriade, M. & Contreras, D. Spike-Wave Complexes and Fast Components of Cortically Generated Seizures. I. Role of Neocortex and Thalamus. J. Neurophysiol. 80, 1439–1455 (1998).

17. Behrens, T. E. J. et al. Non-invasive mapping of connections between human thalamus and cortex using diffusion imaging. Nat. Neurosci. 6, 750–757 (2003).

18. Penfield, W. Epileptic automatism and the centrencephalic integrating system. Res. Publ. - Assoc. Res. Nerv. Ment. Dis. 30, 513–528 (1952).

19. Arthuis, M. et al. Impaired consciousness during temporal lobe seizures is related to increased long-distance cortical–subcortical synchronization. Brain 132, 2091–2101 (2009).

20. Zhang, Y. et al. Thalamocortical structural connectivity abnormalities in drug-resistant generalized epilepsy: A diffusion tensor imaging study. Brain Res. 1727, 146558 (2020).

21. Salanova, V. et al. Long-term efficacy and safety of thalamic stimulation for drug-resistant partial epilepsy. Neurology 84, 1017–1025 (2015).

22. Cukiert, A., Cukiert, C. M., Burattini, J. A. & Mariani, P. P. Seizure outcome during bilateral, continuous, thalamic centromedian nuclei deep brain stimulation in patients with generalized epilepsy: a prospective, open-label study. Seizure 81, 304–309 (2020).

23. Valentín, A. et al. Deep brain stimulation of the centromedian thalamic nucleus for the treatment of generalized and frontal epilepsies. Epilepsia 54, 1823–1833 (2013).

24. Cooper, I. S. et al. Evoked metabolic responses in the limbic-striate system produced by stimulation of anterior thalamic nucleus in man. Int. J. Neurol. 18, 179–187 (1984).

25. Kaufmann, E. et al. Long-term evaluation of anterior thalamic deep brain stimulation for epilepsy in the European MORE registry. Epilepsia 65, 2438–2458 (2024).

26. Salanova, V. et al. The SANTÉ study at 10 years of follow-up: Effectiveness, safety, and sudden unexpected death in epilepsy. Epilepsia 62, 1306–1317 (2021).

27. Weitz, A. J. et al. Optogenetic fMRI reveals distinct, frequency-dependent networks recruited by dorsal and intermediate hippocampus stimulations. NeuroImage 107, 229–241 (2015).

28. Bhattacharyya, K. B. James Wenceslaus Papez, His Circuit, and Emotion. Ann. Indian Acad. Neurol. 20, 207–210 (2017).

29. Guye, M. et al. The role of corticothalamic coupling in human temporal lobe epilepsy. Brain J. Neurol. 129, 1917–1928 (2006).

30. Capecchi, F., Mothersill, I. & Imbach, L. L. The medial pulvinar as a subcortical relay in temporal lobe status epilepticus. Seizure 81, 276–279 (2020).

31. Pizzo, F. et al. The Ictal Signature of Thalamus and Basal Ganglia in Focal Epilepsy. Neurology 96, e280–e293 (2021).

32. Rosenberg, D. S. et al. Involvement of Medial Pulvinar Thalamic Nucleus in Human Temporal Lobe Seizures. Epilepsia 47, 98–107 (2006).

33. Ikegaya, N. et al. Thalamic stereoelectroencephalography for neuromodulation target selection: Proof of concept and review of literature of pulvinar direct electrical stimulation. Epilepsia 65, e79–e86 (2024).

34. Burdette, D., Mirro, E. A., Lawrence, M. & Patra, S. E. Brain-responsive corticothalamic stimulation in the pulvinar nucleus for the treatment of regional neocortical epilepsy: A case series. Epilepsia Open 6, 611–617 (2021).

35. Vilela-Filho, O. et al. Ventral intermediate nucleus deep brain stimulation for treatment-resistant focal aware motor seizures: illustrative case. J. Neurosurg. Case Lessons 5, CASE2320 (2023).

36. Cooper, I. S., Amin, I. & Gilman, S. The effect of chronic cerebellar stimulation upon epilepsy in man. Trans. Am. Neurol. Assoc. 98, 192–196 (1973).

37. Ilinsky, I. et al. Human Motor Thalamus Reconstructed in 3D from Continuous Sagittal Sections with Identified Subcortical Afferent Territories. eNeuro 5, ENEURO.0060-18.2018 (2018).

38. Colombet, B., Woodman, M., Badier, J. M. & Bénar, C. G. AnyWave: a cross-platform and modular software for visualizing and processing electrophysiological signals. J. Neurosci. Methods 242, 118–126 (2015).

39. Granger, C. W. J. Investigating Causal Relations by Econometric Models and Cross-spectral Methods. Econometrica 37, 424–438 (1969).

40. Milosevic, L. et al. Physiological mechanisms of thalamic ventral intermediate nucleus stimulation for tremor suppression. Brain 141, 2142–2155 (2018).

41. Roehri, N., Lina, J.-M., Mosher, J. C., Bartolomei, F. & Benar, C.-G. Time-Frequency Strategies for Increasing High-Frequency Oscillation Detectability in Intracerebral EEG. IEEE Trans. Biomed. Eng. 63, 2595–2606 (2016).

42. Roehri, N., Pizzo, F., Bartolomei, F., Wendling, F. & Bénar, C.-G. What are the assets and weaknesses of HFO detectors? A benchmark framework based on realistic simulations. PLOS ONE 12, e0174702 (2017).

43. Velasco, F. et al. Centromedian Nucleus and Epilepsy. J. Clin. Neurophysiol. 38, 485 (2021).

44. Yamada, K. et al. Somatotopic Organization of Thalamocortical Projection Fibers as Assessed with MR Tractography. Radiology 242, 840–845 (2007).

45. Jbabdi, S., Woolrich, M. W. & Behrens, T. E. J. Multiple-subjects connectivity-based parcellation using hierarchical Dirichlet process mixture models. NeuroImage 44, 373–384 (2009).

46. Aupy, J. et al. Cortico-striatal synchronization in human focal seizures. Brain 142, 1282–1295 (2019).

47. Lopes da Silva, F., Pijn, J. P. & Boeijinga, P. Interdependence of EEG signals: linear vs. nonlinear associations and the significance of time delays and phase shifts. Brain Topogr. 2, 9–18 (1989).

48. Bartolomei, F., Wendling, F., Bellanger, J.-J., Régis, J. & Chauvel, P. Neural networks involving the medial temporal structures in temporal lobe epilepsy. Clin. Neurophysiol. 112, 1746–1760 (2001).

49. Bertram, E. H., Mangan, P. S., Zhang, D., Scott, C. A. & Williamson, J. M. The Midline Thalamus: Alterations and a Potential Role in Limbic Epilepsy. Epilepsia 42, 967–978 (2001).

50. Cassidy, R. M. & Gale, K. Mediodorsal thalamus plays a critical role in the development of limbic motor seizures. J. Neurosci. Off. J. Soc. Neurosci. 18, 9002–9009 (1998).

51. Evangelista, E. et al. Does the Thalamo-Cortical Synchrony Play a Role in Seizure Termination? Front. Neurol. 6, (2015).

52. Soulier, H. et al. The anterior and pulvinar thalamic nuclei interactions in mesial temporal lobe seizure networks. Clin. Neurophysiol. 150, 176–183 (2023).

53. Vetkas, A. et al. Deep brain stimulation targets in epilepsy: Systematic review and meta-analysis of anterior and centromedian thalamic nuclei and hippocampus. Epilepsia 63, 513–524 (2022).

54. Krauss, J. K. et al. Technology of deep brain stimulation: current status and future directions. Nat. Rev. Neurol. 17, 75–87 (2021).

55. Agashe, S. et al. Centromedian Nucleus of the Thalamus Deep Brain Stimulation for Genetic Generalized Epilepsy: A Case Report and Review of Literature. Front. Hum. Neurosci. 16, (2022).

56. Graves, N. M. & Fisher, R. S. Chapter 16 Neurostimulation for Epilepsy, Including a Pilot Study of Anterior Nucleus Stimulation. Neurosurgery 52, 127 (2005).

57. Kim, S. H. et al. Long-term follow-up of anterior thalamic deep brain stimulation in epilepsy: A 11-year, single center experience. Seizure 52, 154–161 (2017).

58. Guo, W. et al. Defining the optimal target for anterior thalamic deep brain stimulation in patients with drug-refractory epilepsy. J. Neurosurg. 134, 1054–1063 (2021).

59. Oh, Y.-S. et al. Cognitive improvement after long-term electrical stimulation of bilateral anterior thalamic nucleus in refractory epilepsy patients. Seizure 21, 183–187 (2012).

60. Park, H. R. et al. The Role of Anterior Thalamic Deep Brain Stimulation as an Alternative Therapy in Patients with Previously Failed Vagus Nerve Stimulation for Refractory Epilepsy. Stereotact. Funct. Neurosurg. 97, 176–182 (2019).

61. Norden, A. D. & Blumenfeld, H. The role of subcortical structures in human epilepsy. Epilepsy Behav. 3, 219–231 (2002).

62. Avoli, M. A brief history on the oscillating roles of thalamus and cortex in absence seizures. Epilepsia 53, 779–789 (2012).

63. W, P. Highest level seizures. Res Publ Nerv Ment Dis 26, 252–271 (1947).

64. Hwang, K., Bertolero, M. A., Liu, W. B. & D’Esposito, M. The Human Thalamus Is an Integrative Hub for Functional Brain Networks. J. Neurosci. 37, 5594–5607 (2017).

65. Wu, T. Q. et al. Multisite thalamic recordings to characterize seizure propagation in the human brain. Brain 146, 2792–2802 (2023).

66. Aracri, P., de Curtis, M., Forcaia, G. & Uva, L. Enhanced thalamo-hippocampal synchronization during focal limbic seizures. Epilepsia 59, 1774–1784 (2018).

67. Destexhe, A., Contreras, D. & Steriade, M. Mechanisms underlying the synchronizing action of corticothalamic feedback through inhibition of thalamic relay cells. J. Neurophysiol. 79, 999–1016 (1998).

68. Schindler, K., Leung, H., Elger, C. E. & Lehnertz, K. Assessing seizure dynamics by analysing the correlation structure of multichannel intracranial EEG. Brain 130, 65–77 (2007).

69. Aiello, G. et al. Functional network dynamics between the anterior thalamus and the cortex in deep brain stimulation for epilepsy. Brain 146, 4717–4735 (2023).

70. Montgomery, E. B. & Gale, J. T. Mechanisms of action of deep brain stimulation(DBS). Neurosci. Biobehav. Rev. 32, 388–407 (2008).

71. Boëx, C., Vulliémoz, S., Spinelli, L., Pollo, C. & Seeck, M. High and low frequency electrical stimulation in non-lesional temporal lobe epilepsy. Seizure 16, 664–669 (2007).

72. Wyckhuys, T., Raedt, R., Vonck, K., Wadman, W. & Boon, P. Comparison of hippocampal Deep Brain Stimulation with high (130 Hz) and low frequency (5 Hz) on afterdischarges in kindled rats. Epilepsy Res. 88, 239–246 (2010).

