## Extended Data Figures for "Personalized Thalamic Electrical Stimulation for Focal Epilepsy"

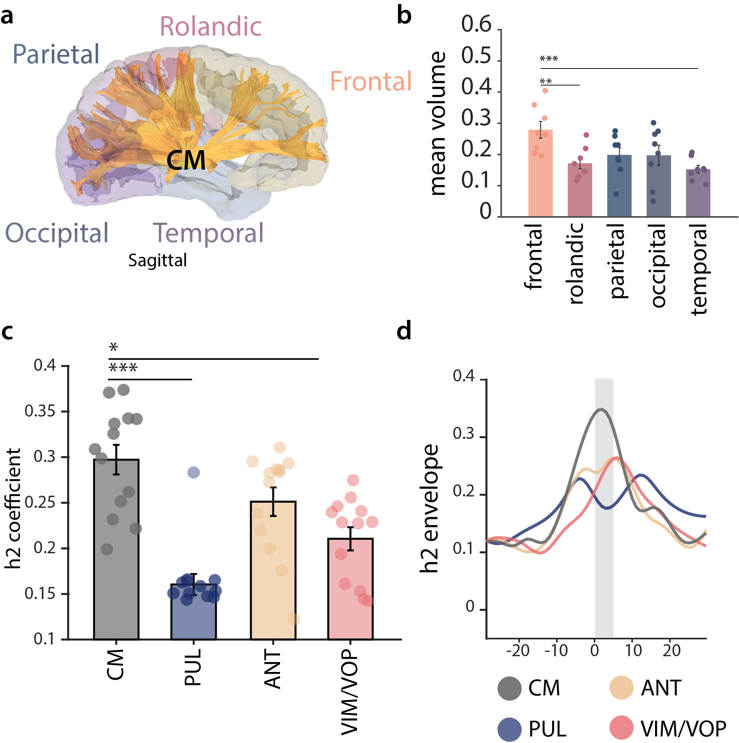

**Extended Data Figure 1: CM nucleus connectivity for generalized epilepsy |** (**a-b**): structural connectivity of the CM. (**a**): representative example (S01) of fiber tracts of the CM. (**b**): Volume of thalamocortical projections (mean ± standard error over 8 patients) from CM to each cortical lobe normalized by the total volume of fibers. (**c-d**): functional connectivity of the CM in generalized epilepsy. (**c**): mean h2 coefficient averaged across 24 seizures, for n=13 bipolar channels and the CM, PUL, ANT and VIM/VOP. (**d**): example of h2 coefficient between the CM, ANT, PUL and VIM/VOP and an epileptogenic zone contact over the course of a seizure. For visualization, we report the h2 envelope to reduce noise.

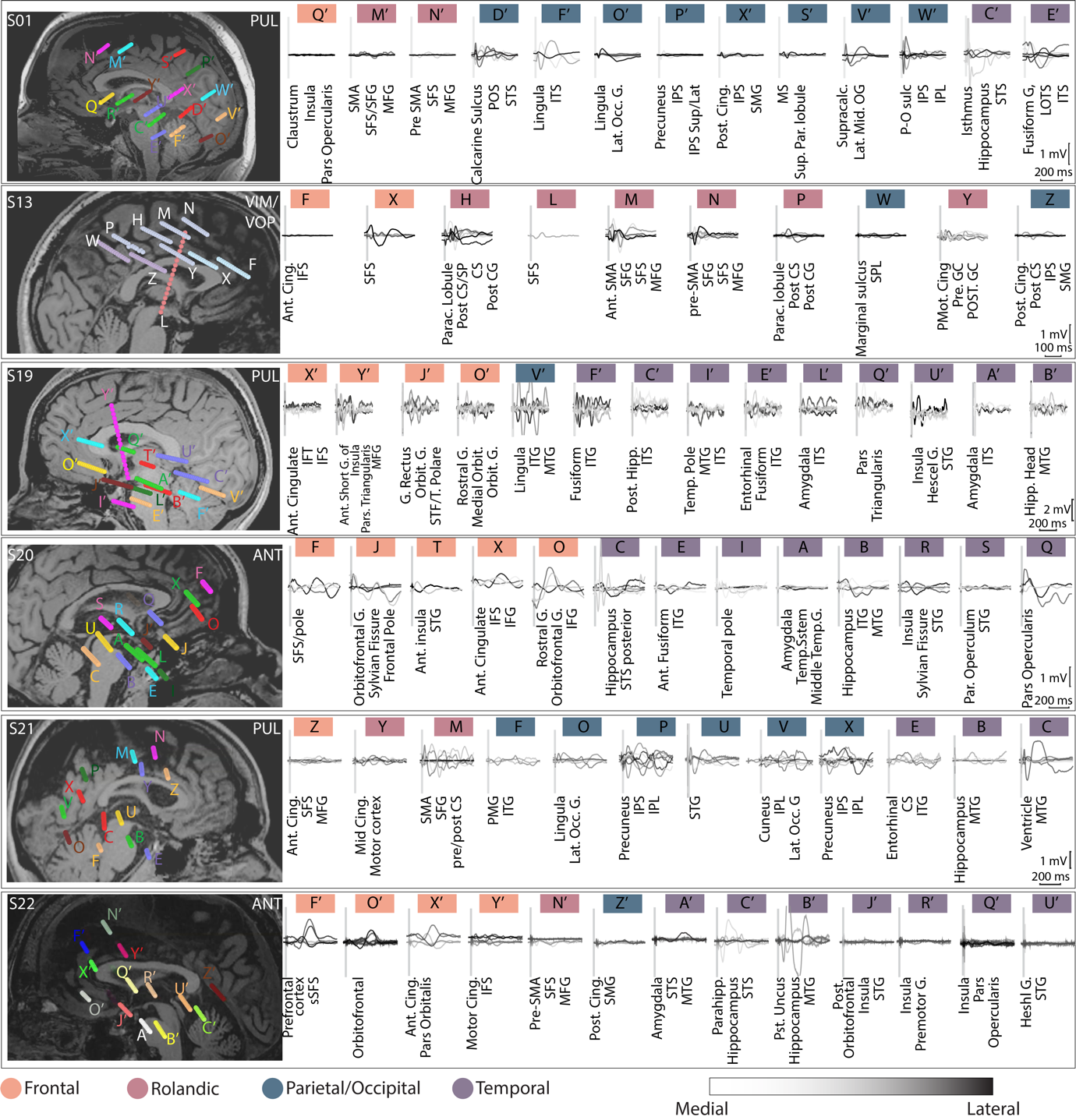

### **Extended Data Figure 2: Thalamocortical evoked potentials in all subjects**

For each patient, we report the SEEG localization overlapped with the MRI (*left*). Stimulation triggered averages for each contact, color coded as medial to lateral (light gray to black) and brain structures involved below each plot (*right*). Abbreviations: ITG=inferior temporal gyrus, IFG = inferior frontal gyrus, MTG = middle temporal gyrus, MFG = middle frontal gyrus, STS/STG = superior temporal sulcus/gyrus, SFS = superior frontal sulcus, G = gyrus, S = sulcus, Par = parietal, Temp = temporal, Hipp = hippocampal, Ant = anterior, Cing = cingulate, Sup = superior, SMA = supplementary motor area, IPL = inferior parietal lobe, POS = parieto-occipital sulcus, IPS = intraparietal sulcus.

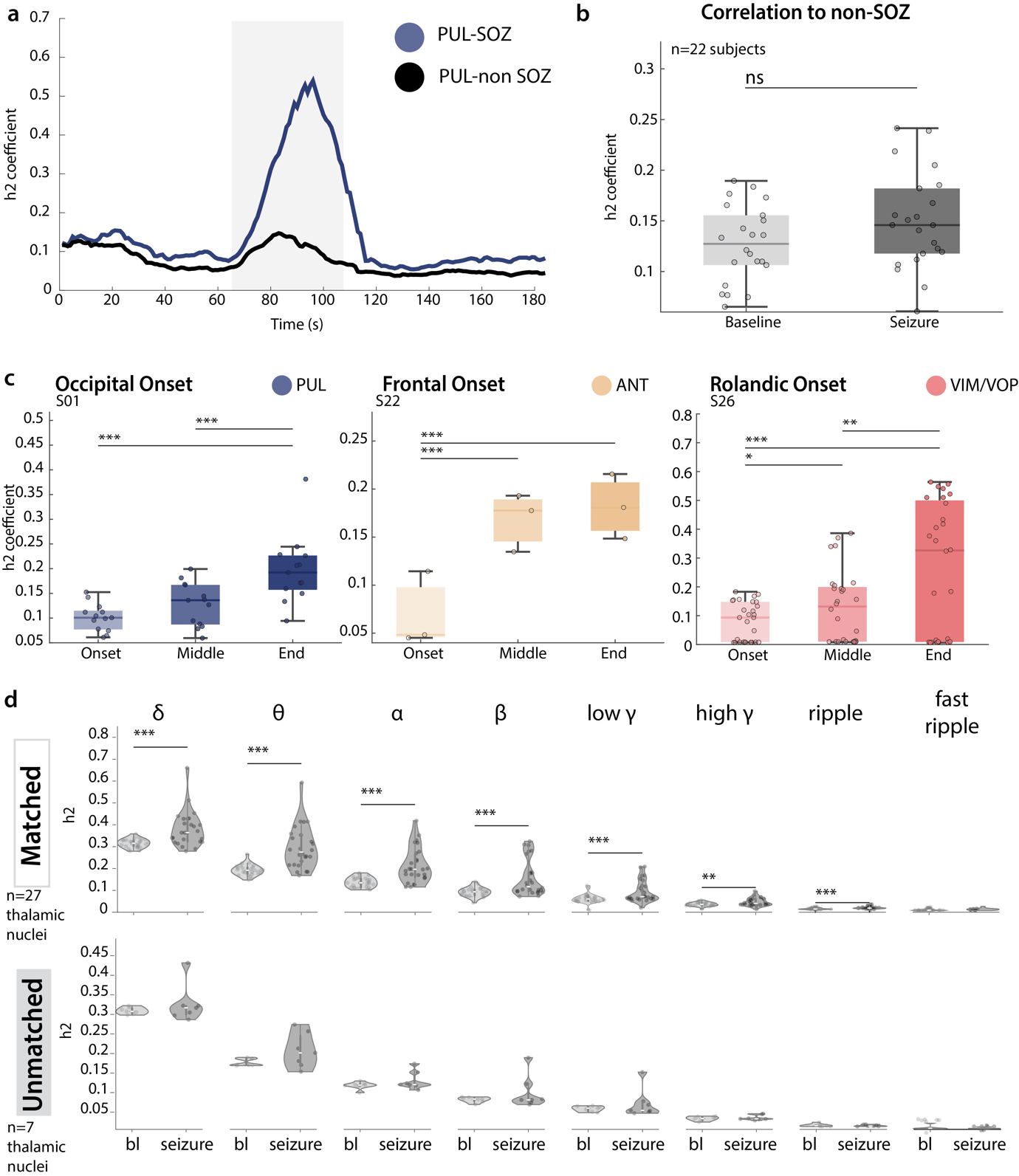
**Extended Data Figure 3: Thalamocortical synchronization with non-linear correlation coefficient|** (**a-b**) h2 correlation of thalamus and non-SOZ areas. (**a**): representative example of the h2 coefficient during the course of a seizure of the PUL with a matched SOZ (blu) and a non-SOZ (black) cortical region in S03. (**b**) Boxplot of the mean h2 coefficient between each nucleus and the mean of 3 non-SOZ contacts during seizures. (**c**): analysis of h2 coefficient during multiple phases of the seizure (onset, middle and termination). One representative patient for each nucleus (PUL, ANT, VIM/VOP) is represented (n=13, n=3, n=28 seizures respectively). (**d**) Violin plot of the mean h2 coefficient during the seizure for all matched (n=27 nuclei) and unmatched (n=7 nuclei) thalami-SOZ pairs, for different frequency bands. Each point represents the mean across all the seizures.

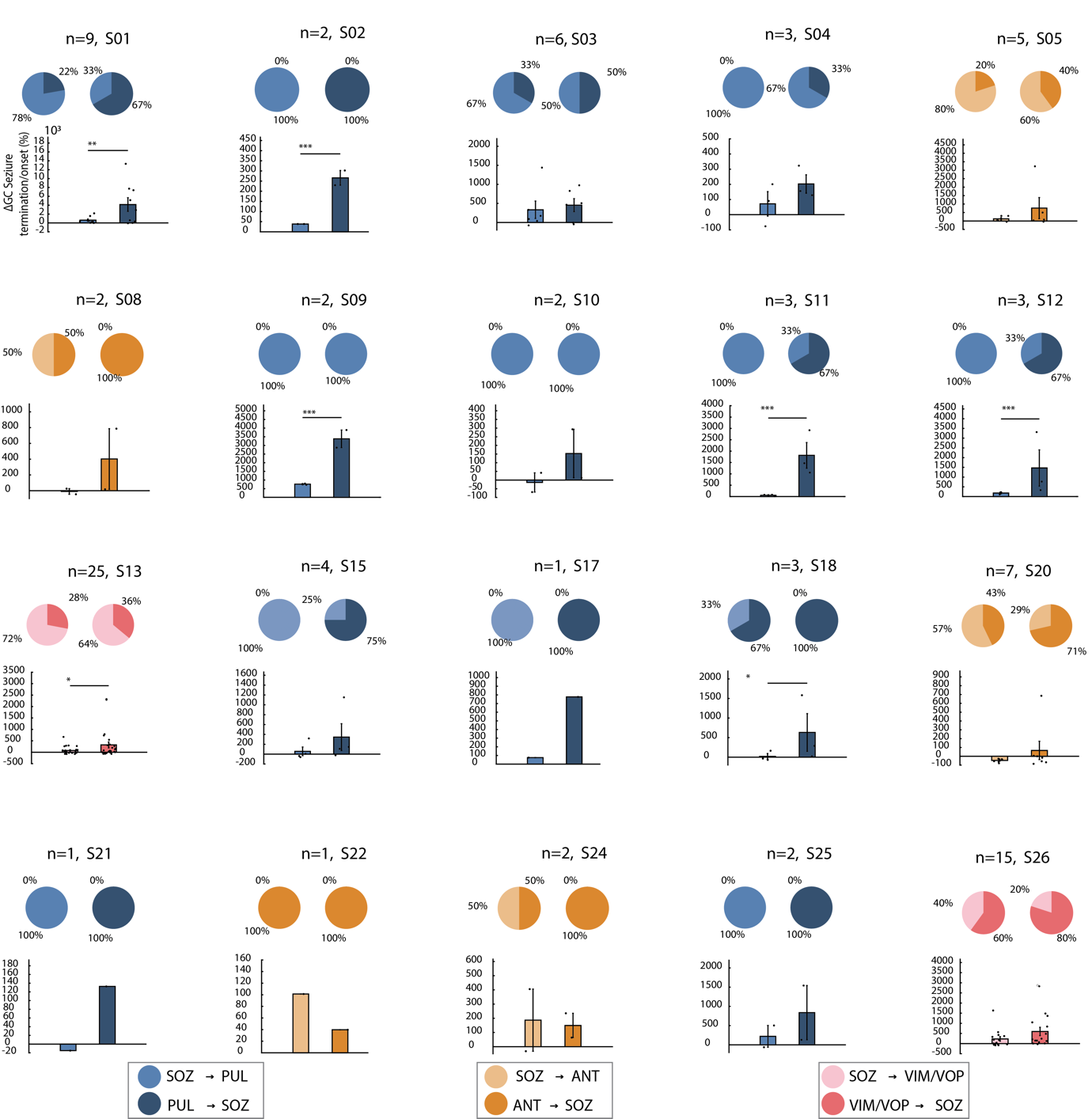

### **Extended Data Figure 4: Single subject Granger Causality results|** For each subject, analysis of: Pie charts illustrating the proportion of seizures with higher corticothalamic (SOZ→th, light color) or thalamocortical (th→SOZ, dark color GC coefficient for the initiation (left) and termination (right) epoch. (top); Bar plot of the relative percentage change of GC coefficient from initiation to termination. Each data point represents the value of an individual seizure (bottom). Each subject title reports the number of seizures.

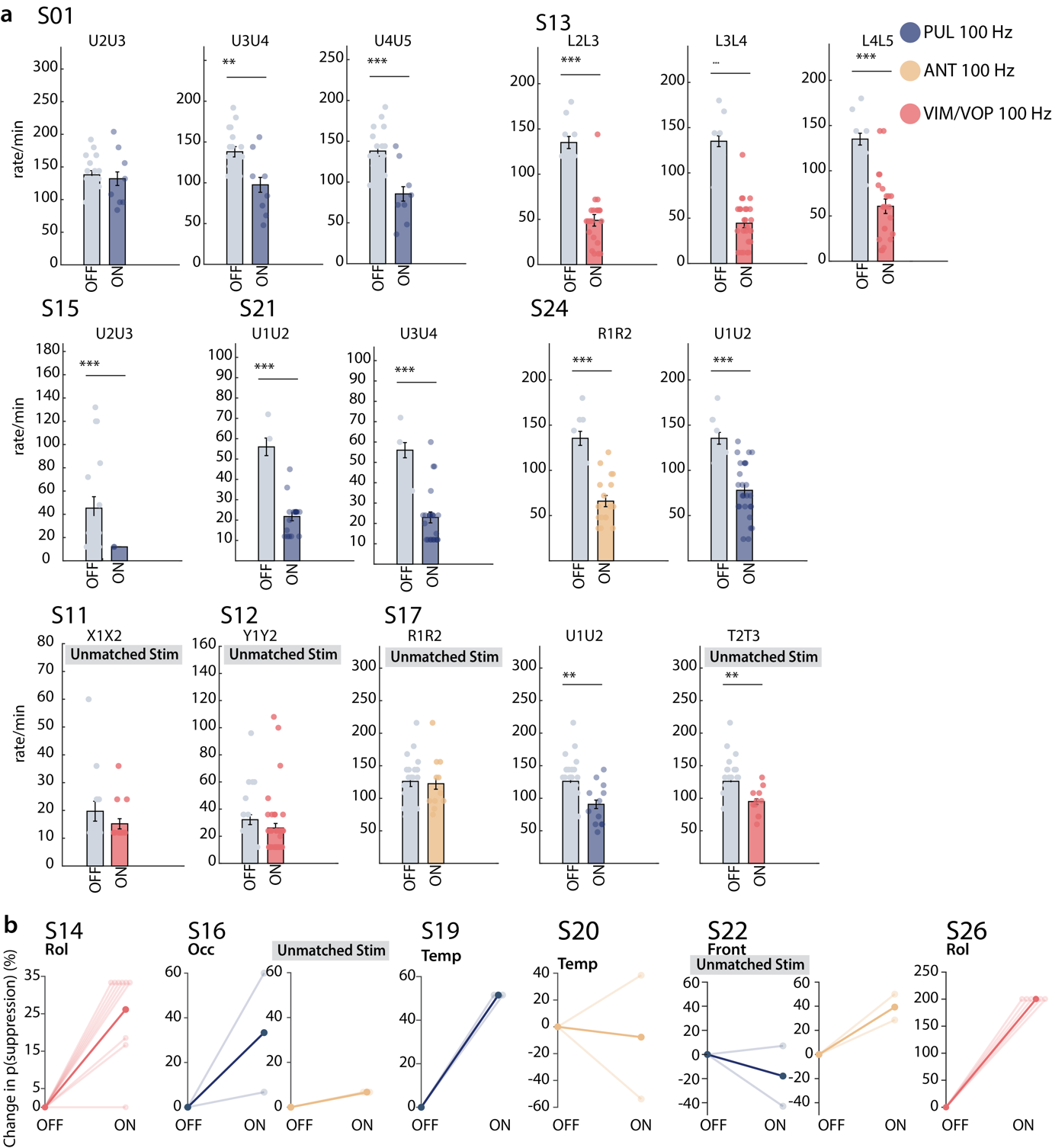

### **Extended Data Figure 5: Stimulation immediate effect |** (**a**): bar plot of the IED rate for each pair of bipolar contacts (explicated in the title) stimulated in each nucleus. (**b**): For all subjects, percentage change relative to baseline of the probability of IEDs suppression in matched (left) and unmatched (right) stimulation. Bright lines represent different stimulation trials while dark lines represent the mean.
