## Supplementary Tables for "Personalized Thalamic Electrical Stimulation for Focal Epilepsy"

| **Subject N** | **Gender** | **Age at surgery** | **Duration of epilepsy** | | **Type of seizure/semiology** | | **SOZ** |
| --- | --- | --- | --- | --- | --- | --- | --- |
| **1** | M | 30-35 | 21 | CPS + GTC | | Occipital | |
| **2** | M | 40-45 | 42.5 | SPS + GTC | | Temporal | |
| **3** | F | 35-39 | 28 | SPS + GTC | | Parietal | |
| **4** | M | 30-35 | 31 | SPS + GTC | | Temporal | |
| **5** | M | 25-29 | 7 | SPS + GTC | | Frontal | |
| **6** | F | 45-49 | 44 | SPS + CPS | | Temporal | |
| **7** | M | 20-25 | 16 | SPS + CPS | | Temporal | |
| **8** | F | 40-45 | 30 | SPS + CPS + GTC | | Temporal | |
| **9** | M | 35-39 | 21 | SPS + GTC | | Temporal | |
| **10** | M | 20-25 | 10 | SPS + CPS + GTC | | Temporal | |
| **11** | M | 35-39 | 4 | SPS + CPS + GTC | | Temporal | |
| **12** | M | 20-25 | 1 | CPS + GTC | | Temporal | |
| **13** | M | 40-45 | 26 | SPS + GTC | | Rolandic | |
| **14** | M | 50-55 | 12 | SPS + CPC | | Rolandic | |
| **15** | F | 60-65 | 2 | CPS + GTC | | Temporal | |
| **16** | M | 30-35 | 32.5 | SPS + GTC | | Parietal/Occipital | |
| **17** | F | 20-25 | 11 | SPS + GTC | | Parietal | |
| **18** | M | 20-25 | 5 | SPS + CPS | | Temporal | |
| **19** | M | 25-29 | 7 | SPS + CPS | | Temporal | |
| **20** | F | 35-39 | 10 | SPS + GTC | | Temporal | |
| **21** | M | 25-29 | 28 | CPS | | Occipital | |
| **22** | M | 60-65 | 51 | CPS + GTC | | Frontal | |
| **23** | M | 35-39 | 25 | SPS + GTC | | Rolandic | |
| **24** | M | 35-39 | 24 | SPS + CPS + GTC | | Temporal | |
| **25** | M | 20-25 | 22 | SPS + CPS + GTC | | Temporal | |
| **26** | M | 15-19 | 4 | CPS + GTC | | Rolandic | |
| **27** | F | 25-29 | 27 | CPS + GTC | | Occipital/Parietal | |
| **28** | F | 40-45 | 8 | CPS | | Occipital/Parietal | |
| **29** | F | 30-35 | 31 | CPS | | Occipital/Parietal | |
| **30** | M | 30-35 | 21 | CPS + GTC | | Occipital | |
| **31** | F | 20-25 | 18 | CPS + GTC | | Occipital | |
| **32** | M | 30-35 | 32 | CPS + GTC | | Frontal | |

**Supplementary Table 1 |** Patient demographic and epilepsy type (SPS = simple partial seizure, CPS = complex partial seizure, GTC = general tonic seizure). Gray cells indicate the patients who were chronically implanted with a hodologically-matched neurostimulation system.

| **Subject N** | **Thalamic Coverage** | | | **Spontaneous Seizures** | **DTI** | **EP** | **Acute stimulation** |
| --- | --- | --- | --- | --- | --- | --- | --- |
|  | **PUL** | **ANT** | **VIM/VOP** |  |  |  |  |
| **1** | x | x |  | 13 |  | X | a |
| **2** | x |  |  | 4 |  |  |  |
| **3** | x |  |  | 6 |  |  |  |
| **4** | x | x |  | 7 | X |  |  |
| **5** | x | x |  | 9 |  |  |  |
| **6** | x |  |  | 8 | X |  |  |
| **7** | x |  |  | 10 | X |  |  |
| **8** | x | x |  | 5 | X |  |  |
| **9** | x |  |  | 2 |  |  |  |
| **10** | x |  |  | 2 |  |  |  |
| **11** | x |  | x | 6 |  |  | a |
| **12** | x |  | x | 5 | X |  | a |
| **13** |  |  | x | 51 | X | X | a |
| **14** |  |  | X | 0 |  |  | b |
| **15** | x |  |  | 4 |  |  | a |
| **16** | x | x |  | 0 |  |  | b |
| **17** | x | x | x | 7 |  |  | a |
| **18** | x |  |  | 4 |  |  |  |
| **19** | x |  |  | 0 |  | X | b |
| **20** |  | x |  | 11 | X | X | b |
| **21** | x |  |  | 22 |  | X | a |
| **22** | x | x |  | 3 |  | X | b |
| **23** |  |  | x | 4 |  |  |  |
| **24** | x | x |  | 3 | X |  | a |
| **25** | x |  |  | 2 |  |  |  |
| **26** |  |  | x | 28 |  |  | b |

### **Supplementary Table 2 |** Thalamic coverage and performed experiments for SEEG population (S1-S26)

| **Subject N** | **SOZ categorization** | **SOZ anatomical area** | **non-SOZ area 1** | **non-SOZ area 2** | **non-SOZ area 3** |
| --- | --- | --- | --- | --- | --- |
| **1** | Occipital | Lingula | Superior frontal sulcus (SFS) | SFS/SFG (frontal eye field) | IPS ( sup/med) |
| **2** | Temporal | Mid STG | Amygdala | Head of hippocampus | Post precuneus |
| **3** | Parietal | Parietal encephalomalacia | Infracalcarine | Mesial precentral gyrus | Frontal eye field / anterior SMA |
| **4** | Temporal | Mesial temporal lobe | Basal temporal / occipital lobe | Basal temporal / occipital lobe | Post orbitofrontal |
| **5** | Frontal | Orbitofrontal | Frontal pole lat | Tail of hippocampus | Entorhinal |
| **6** | Temporal | Mesial temporal lobe | Frontal operculum | Junction of precentral sulcus / IFS | Mid frontal gyrus |
| **7** | Temporal | Mesial temporal lobe | Supracalcarine | Infracalcarine | Anterior precuneus |
| **8** | Temporal | Mesial temporal lobe | Frontal pole med | Anterior orbitofrontal | Frontal pole lat |
| **9** | Temporal | Mesial temporal lobe | Piddle temporal gyrus | Parahippocampus | Posterior orbitofrontal |
| **10** | Temporal | Mesial temporal lobe | Dorsal striatum | Post cingulate | Superior temporal sulcus |
| **11** | Temporal | Mesial temporal lobe | - | - | - |
| **12** | Temporal | Mesial temporal lobe | Parahippocampus | Superior temporal gyrus | Middle temporal gyrus |
| **13** | Rolandic | Motor cingulate | Inferior frontal sulcus | Mid frontal gyrus | Anterior SMA |
| **14** | Rolandic | Motor Cortex | - | - | - |
| **15** | Temporal | Mesial temporal lobe | Collateral sulcus | Temporal pole | Dorsal amygdala |
| **16** | Parietal/Occipital | Superior Parietal Lobule/Cuneus | - | - | - |
| **17** | Parietal | Inferior Parietal Lobule | Post cingulate | Precentral leg motor | Superior parietal lobule |
| **18** | Temporal | Superior Temporal Sulcus | Post hippocampus | Anterior hippocampus | Amygdala |
| **19** | Temporal | Mesial temporal lobe | - | - | - |
| **20** | Temporal | Mesial temporal lobe | Superior temporal sulcus | Orbitofrontal | Orbitofrontal gyrus |
| **21** | Occipital | Lateral Occipital Gyrus | Anterior cingulate | Mid cingulate | Anterior cingulate |
| **22** | Frontal | Orbitofrontal | Anterior cingulate | Head of hippocampus | SFG PreSMA |
| **23** | Rolandic | Motor Cingulate/Cortex | Indusium griseum | MFG | Supramarginalis |
| **24** | Temporal | Anterior Uncus | Motor cingulate | Claustrum | Temporal pole |
| **25** | Temporal | Mesial temporal lobe | Pars triangularis | Superior temporal gyrus planum polare | Basal occipital |
| **26** | Rolandic | Motor Cortex | Motor cortex med. | preSMA | Paracentral lobule |

### **Supplementary Table 3 |** Anatomical location of SOZ and non-SOZ regions for SEEG population.

| **Subject N** | **Target** | **Follow-up (months)** | **Stimulation frequency (Hz)** | **Stimulation amplitude (mA)** | **SOZ** |
| --- | --- | --- | --- | --- | --- |
| **16** | PUL | 24 | 125 | 1.8 | Occipital/Parietal |
| **27** | PUL | 12 | 145 | 3 | Occipital/Parietal |
| **28** | PUL | 12 | 130 | 3 | Occipital/Parietal |
| **29** | PUL | 12 | 135 | 2 | Occipital/Parietal |
| **30** | PUL | 9 | 135 | 2.5 | Occipital |
| **31** | PUL | 30 | 130 | 2 | Occipital |
| **32** | ANT | 12 | 145 | 2 | Frontal |

### **Supplementary Table 4 |** Details about chronic participants and chronic implant received.
